# Protein kinase C eta enhances Golgi-localized signaling and is associated with Alzheimer’s disease using a recessive mode of inheritance

**DOI:** 10.1101/2025.05.13.25327562

**Authors:** Maria Celeste Gauron, Dmitry Prokopenko, Sanghun Lee, Sarah A. Wolfe, Julian Hecker, Julian Willett, Mohammad Waqas, Gema Lordén, Yimin Yang, Joshua E. Mayfield, Isabel Castanho, Kristina Mullin, Sarah Morgan, Georg Hahn, Dawn L. Demeo, Winston Hide, Lars Bertram, Christoph Lange, Alexandra C. Newton, Rudolph E. Tanzi

## Abstract

The identification of Alzheimer’s disease (AD)-associated genomic variants has provided powerful insight into disease etiology. Genome-wide association studies (GWAS) for AD have successfully identified new targets but have almost exclusively utilized additive genetic models. Here, we performed a family-based GWAS under a recessive inheritance model using whole genome sequencing from families affected by AD. We found that the variant, rs7161410, located in an intron of the *PRKCH* gene, encoding protein kinase C eta (PKCη), was associated with AD risk (p-value=1.41 × 10−7). Further analysis revealed a rare *PRKCH* missense mutation K65R in linkage disequilibrium with rs7161410, which was present in homozygous carriers of the rs7161410 risk allele. We show that this mutation leads to enhanced localization and signaling of PKCη at the Golgi. The novel genetically-validated association of aberrant PKCη signaling with AD opens avenues for new therapeutic targets aimed at prevention and treatment.

**One Sentence Summary:** Protein kinase C eta enhances Golgi-localized signaling and is associated with Alzheimer’s disease.

## Introduction

Alzheimer’s disease (AD) is the most common form of dementia accounting to 60–80% of cases in the elderly. AD is expected to increase worldwide to 66 million by 2030, and 131 million by 2050^1^. AD is a heterogeneous, neurodegenerative disorder broadly classified into two subtypes: early-onset and late-onset. Early-onset familial AD (EOAD) is generally inherited in an autosomal dominant pattern due to mutations in *APP* (amyloid-b precursor protein), *PSEN1* (presenilin 1), and *PSEN2* (presenilin 2) genes, however there are also cases of EOAD which cannot be linked to any of these three genes. About 90% of EOAD cases from 32 United States Alzheimer’s Disease Centers were reported to be due to autosomal recessive causes^2^. The inheritance pattern of late-onset AD (LOAD) is polygenic, with the *APOE* (Apolipoprotein E) ε4 allele being the strongest genetic risk factor^3^. To date, various genes implicated in AD have been identified by genome wide association studies (GWAS) utilizing an additive model (AM) of inheritance, assuming a uniform and linear increase in risk^4,5^. To broaden our capture of AD-associated risk variants, we performed a family-based GWAS under a recessive model (RM) using whole genome sequencing (WGS) data in order to trace potential recessive loci that confer susceptibility to AD. Using this model, we identified a variant in a PKC gene family member, *PRKCH*, encoding protein kinase C eta (PKCη), as a risk factor for AD. Further analysis led to the identification of rare and highly-penetrant AD-associated variants resulting in missense mutations in *PRKCH* that alter function.

PKCη is a member of the novel class of PKC isozymes, Ser/Thr kinases that are activated by the second messenger diacylglycerol (DG)^6–8^. Novel PKC isozymes are closely related to the conventional PKC isozymes, which are additionally regulated by Ca^2+^. Originally identified in the brain, conventional PKC family members are being increasingly recognized as key players in maintaining normal brain function^9^. These isozymes regulate synapse morphology, receptor turnover, and cytoskeletal integrity. Unbiased phosphoproteomics analyses identify enhanced signaling by PKC as one of the earliest events in the pathology associated with Alzheimer’s Disease^10^. Our previous genetic analyses identified the conventional isozyme PKCα as a driver in the pathology of AD. Specifically, highly penetrant variants of the PKCα gene, *PRKCA*, which conferred increased risk for AD, were shown to be gain-of-function mutations^11^. Introduction of one such variant (PKCα M489V) in a mouse model was sufficient to rewire the brain phosphoproteome, reduce spine density of neurons, enhance Ab-induced synaptic depression, and cause cognitive decline^12^. Additionally, variants in another conventional PKC member, *PRKCG*, encoding Purkinje cell-localized PKCγ, are causative for Spinocerebellar Ataxia Type 14, likely resulting from enhanced basal signaling^13,14^. However, unlike conventional isozymes, whether novel PKCs drive neurodegenerative diseases has not been previously established.

Here, we used a recessive mode of inheritance in a family-based GWAS using WGS data to identify PKCη as a novel potential AD target. Specifically, we identified an intronic variant in *PRKCH*, rs7161410, to be strongly associated with AD risk (p-value=1.41 × 10^−7^). Additionally, we identified five exonic variants in strong linkage disequilibrium (LD) with the risk allele of rs7161410; of these, only one was present in homozygous carriers of rs7161410. This variant corresponds to missense mutation, K65R, in a surface-exposed residue in the C2 domain of PKCη. *In vitro* kinase assays revealed that this mutation does not alter the intrinsic biochemical properties of PKCη. In contrast, cellular studies using FRET-based reporters revealed that the mutation leads to enhanced localization and signaling of PKCη at the Golgi. In summary, performing a family-based GWAS of WGS data, using a RM of inheritance, we identified a novel AD-associated variant in tight LD with a missense mutation in *PRKCH* that alters PKCη function.

## Results

### Identification of *PRKCH* as an AD marker

The AD families included in this study originated from two cohorts: The National Institute of Mental Health (NIMH)^15^ and the family component of the National Institute of Aging AD Sequencing Project (NIA ADSP)^16^. By design, carriers of ε4/ε4 individuals were not included in the sequencing efforts of NIA ADSP. After the quality control filtering, two WGS familial AD cohorts with 1,393 individuals (NIMH; 446 multiplex families) and 854 individuals (NIA; 159 multiplex families) were merged together. In the NIMH cohort, the majority (95.33%) are European American, i.e. 1,328 out of 1,393, and 68.05% of whom are women: 948 subjects with AD (mean [±SD] age at onset or screening, 72 ± 10 years; range 31 to 100), and 445 unaffected subjects. In the ADSP cohort, European American, Caribbean Hispanic, and Dutch ancestry were represented and 63.11% of whom are women: 543 subjects with AD (mean [±SD] age at onset or screening, 72 ± 10 years; range 30 to 90), 296 unaffected subjects, and 15 subjects with an unknown phenotype. Among total a 1,509 of AD subjects from 605 families, the number of EOAD (onset age < 65) and LOAD (onset age ≥ 65) were 246 and 1,263 respectively.

Genome-wide family-based association test (FBAT) outcomes under RM were shown in quantile–quantile and Manhattan plots without evidence of spurious inflation of association test statistics (λ = 1.00, Figure 1a and 1c). The peak association originated on chromosome 19 is approximately 500 bases downstream (3′) of *APOC1* (Apo lipoprotein C1) at 19q13.32. The top single nucleotide polymorphisms (SNPs), rs4420638 and rs56131196 with a *p*_rec_ value of 3.48 × 10^−11^, were much more significant under AM (*p*_add_ = 4.44 × 10^−15^, *p*_add_ = 6.66 × 10^−15^, respectively; Figure 1d and Supplementary Table 1). We also confirmed that *rs429358* (coding for ε4 allele) in *APOE* shows a recessive signal implying a strong AD effect in carriers of the ε4/ε4 genotype (Family size: 54 in RM vs. 144 in AM and *p*_rec_ = 4.18 × 10^−9^vs. *p*_add_ = 1.11 × 10^−15^).

**Figure 1:**
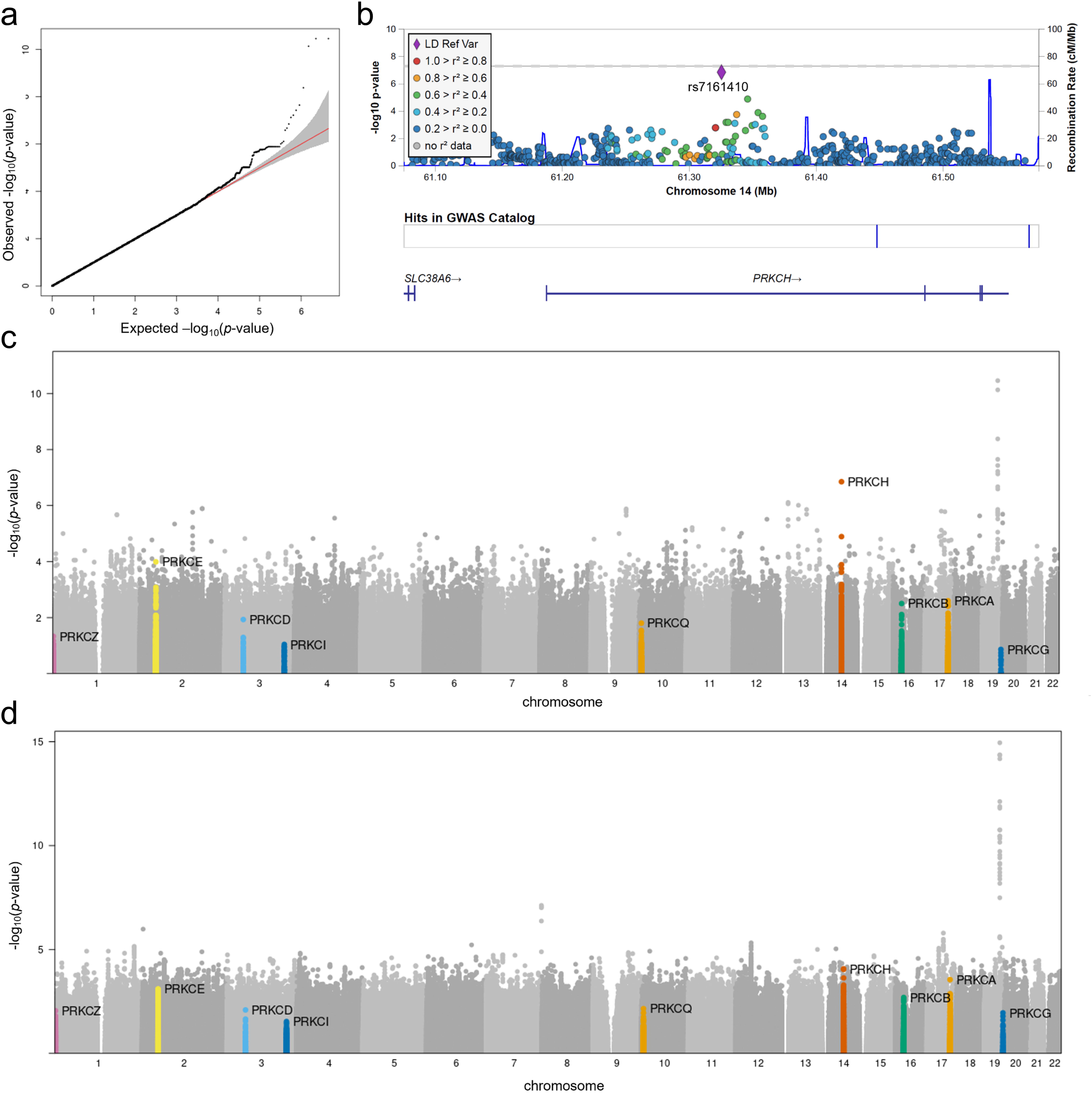
Identification of *PRKCH* as an AD marker. GWAS under recessive model with 2,247 subjects from 558 family’s trios according to AD affection status, shown in Quantile-Quantile plot **(a)** regional plot of the significant variant, rs7161410 on *PRKCH* **(b)**, and Manhattan plot **(c)**, while GWAS under additive model was shown in Manhattan plot **(d)** demonstrating that the hit (rs7161410) on *PRKCH* loses its significance compared to recessive model. Protein kinase C family members were highlighted in each Manhattan plot.

Among the loci exhibiting suggestive genome-wide significance (*p* < 5 × 10^−7^) under RM, only rs7161410, an intronic variant of *PRKCH* (encoding PKCη) in chromosome 14 was more significant compared to AM (*p*_rec_ = 1.41 × 10^−7^ vs. *p*_add_ = 9.20 × 10^−5^, respectively; Figure 1c and 1d, Supplementary Table 1) suggesting that this signal is stronger under a recessive inheritance model. Moreover, the number of informative families was much smaller under RM compared to AM (36 vs. 139, respectively). Despite the considerable discrepancy in size, the p-value is notably more significant in RM, indicating a strong recessive signal. Interestingly, the stratified analysis by ancestry revealed that the signal was the strongest with small informative family sizes in Dominican Families (n=412, number of informative families = 10, and *p*_rec_ = 7.81 × 10^−7^, Supplementary Table 2) while it was significant in non-Hispanic white except Dominican families (n=1,717, number of informative families = 24, and *p*_rec_ = 3.72 × 10^−3^, Supplementary Table 2). However, it was not significant in African Americans (n=84, number of informative families = 2, and *p*_rec_ = 0.617, Supplementary Table 2). The FBAT interaction test (FBAT-GE)^17,18^ showed a significant interaction (p=0.022) between the SNP and a binary indicator, whether or not a subject belongs to the Dominican Families subgroup.

While this allele exhibited association with AD in a family-based WGS sample in Dominican and non-Hispanic white families, we were unable to replicate this recessive association signal in an independent NIA ADSP case-control dataset, *p*_rec_ = 0.89 in African American subpopulation (n=4263), *p*_rec_ = 0.27 in non-Hispanic white subpopulation (n=9609), *p*_rec_ = 0.65 in Hispanic subpopulation (n=8466), and *p*_rec_ = 0.58 in Asian subpopulation (n=2543) and in two biobanks with an AD-by-proxy phenotype (UK Biobank, p_rec_=0.57; AllofUS, p_rec_=0.25).

Next, we set out to identify exonic variants exhibiting potential functional effects, which could, in part, explain the recessive association of the intronic *PRKCH* variant with familial AD. Five SNPs were found to be in tight LD (LD, D’>0.9) with the rs7161410 risk allele, all having predicted moderate or high impact on *PRKCH* (Figure 2a). These corresponded to A19V and K65R in the C2 domain, R149Q in the C2-C1A linker, and V374I and A410S in the kinase domain (Figure 2b). Among these variants, one missense mutation (K65R, *p*_add_=0.074, p_rec_=NA due to no homozygous carriers), which was present only in 8 affected heterozygous carriers in the NIMH AD families was of particular interest. It was the only functional mutation in *PRKCH*, for which, carriers were also homozygous for the minor allele of rs7161410 in both datasets - NIA ADSP unrelated subjects and NIMH and NIA ADSP families - partially accounting for the observed recessive association signal with rs7131410 (Supplementary Table 3 and Supplementary Figure 1). The removal of K65R carriers and their families from the dataset lead to a slight decrease in recessive signal of rs7161410 due to removal of one informative non-Hispanic white family, which contributed to the FBAT test statistic of rs7161410 (Supplementary Table 2). In addition, the K65R was replicated in the latest release of NIA ADSP (v9) with unrelated cases and controls (*p*_add_=0.030, p_rec_=NA due to no homozygous carriers) in which 9 carriers of K65R are homozygous for rs7161410. Among other functional mutations V374I had 39 carriers which were homozygous for the minor allele of rs7161410 in NIA ADSP unrelated subjects but not in NIMH and NIA ADSP families. Mapping of the five mutations onto the modeled structure of PKCη^19^ revealed these are all surface-exposed residues (Figure 2c). Gene-based analysis including the five functional mutations demonstrated significant association in the SKAT/variance-component based region tests (*p*_add_=0.0175).

**Figure 2:**
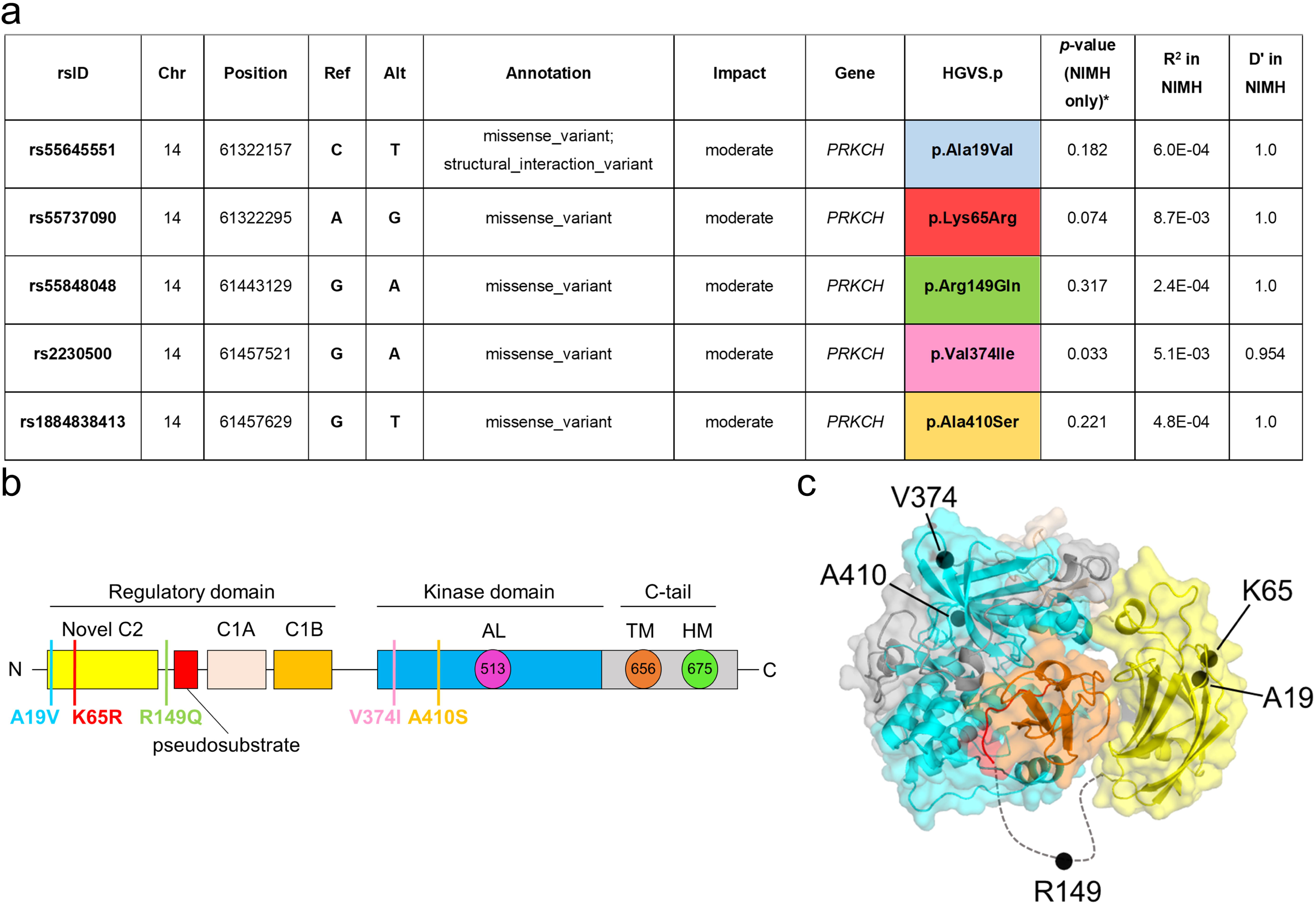
PKCη AD-associated variants. **(a)** Single nucleotide variants in linkage disequilibrium (LD, D’>0.9) with rs7161410 with a moderate or high functional impact. **(b)** Primary structure of PKCη showing domain composition and position of AD-associated variants; novel C2 domain (yellow), autoinhibitory pseudosubstrate (red), C1A domain (tan), diacylglycerol-sensing C1B domain (orange), kinase domain (cyan), and C-terminal tail (grey). Processing phosphorylation sites at the activation loop (Thr513), turn motif (Thr656) and hydrophobic motif (Ser675) are indicated. **(c)** Model for autoinhibited conformation of the novel PKCη isozyme^19^ showing position of AD variants; residues are all surface-exposed.

### Reduced cytosolic activity of AD-associated PKC**η** mutations

To assess whether these AD-associated mutations affect PKCη function, we first addressed their effect on the basal and agonist-evoked activity of PKCη. We used the genetically encoded cytosolic fluorescence resonance energy transfer (FRET)-based biosensor C Kinase Activity Reporter 2 (CKAR2) to monitor PKC activity in the cytosol^20^. COS7 cells co-expressing CKAR2 together with mCherry-tagged PKCη wild-type (WT) or each of the five AD-variants: A19V, K65R, R149Q, V374I and A410S, were treated 1] directly with the PKC inhibitor, Gö6983, to abolish PKC activity (Figure 3a) or 2] first with uridine-5’-triphosphate (UTP), which activates purinergic receptors to elevate DG (and Ca^2+^) to transiently activate PKC, followed by treatment with Gö6983 to abolish its activity (Figure 3c). Cells expressing similar levels of mCherry PKC were selected for analysis. Unlike conventional PKC isozymes, novel PKC isozymes have high basal signaling in the absence of stimulation, which can be assessed by measuring the reduction in FRET upon PKC inhibition. The addition of Gö6983 to cells transfected with the mCherry empty vector to study the activity of endogenous PKCs caused no significant change in FRET (Figure 3a, grey trace), this is because the main cytosolic activity in COS7 cells is driven by conventional PKCs, which are tightly autoinhibited in the absence of agonist stimulation. In contrast, inhibitor treatment caused a significant reduction in FRET ratio in cells overexpressing PKCη WT (Figure 3a, blue trace), reflecting its high basal activity in the absence of agonists. When analyzing the AD-associated mutants, it was observed that each of the five variants had significantly lower basal activity than PKCη WT (Figure 3a and 3b). These data reveal that the AD-associated mutants have lower basal signaling assessed using the cytosolic biosensor compared with PKCη WT.

**Figure 3:**
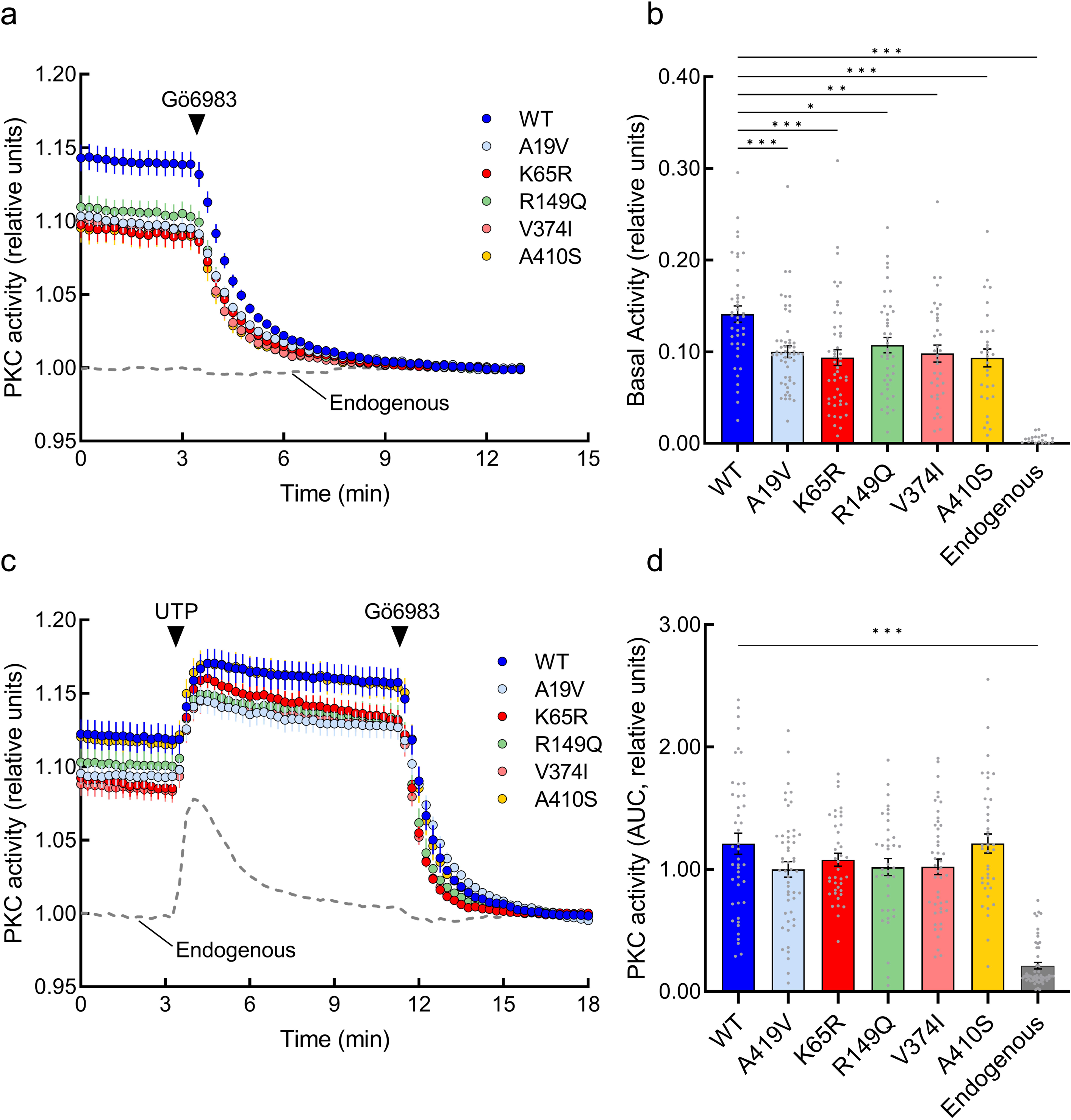
AD-associated PKCη mutations have reduced activity in the cytosol. **(a)** Normalized FRET ratios representing PKC activity in COS7 cells co-expressing the indicated mCherry-tagged PKCη (WT, blue trace; A19V, cyan trace; K65R, red trace; R149Q, green trace; V374I, pink trace; A410S, yellow trace), or mCherry empty vector (Endogenous PKCs, grey trace) and the PKC activity reporter, cytosolic CKAR2. After establishing a baseline for 3 minutes, cells were treated with 1 μM PKC inhibitor Gö6983, indicated by the black arrow. Data are representative of 30-53 cells per condition, from three independent experiments. **(b)** Relative basal activity, quantified using the traces in A by subtraction of the average of the last 2.5 minutes minus the average of the baseline registered pre-inhibitor treatment. Data are the mean ± SEM of three independent experiments, ***p ≤ 0.0001, **p ≤ 0.001, *p ≤ 0.05. **(c)** Normalized FRET ratios of PKC activity in COS7 cells co-expressing the indicated mCherry-tagged PKCη, or mCherry empty vector and CKAR2. After establishing a baseline for 3 minutes, PKCs were activated with 100 μM uridine 5′-triphosphate (UTP), after plateau cells were treated with 1μM Gö6983. Data are representative of 38-52 cells per condition, from four independent experiments. **(d)** Relative PKC activity, quantified as area under the curve (AUC) from 3 min to 11 min as a measure of signaling output. Data are the mean ± SEM of four independent experiments, ***p ≤ 0.0001, **p ≤ 0.001, *p ≤ 0.05.

We next sought to analyze the agonist-evoked activity of PKCη WT compared to the rare variants. Cells co-expressing cytosolic CKAR2 and mCherry-PKCη WT or each one of the variants were treated with UTP to stimulate DG-dependent activity and then treated with Gö6983 to abolish activity (Figure 3c). UTP treatment caused an increase in the activity of endogenous PKC which rapidly decayed to baseline levels, consistent with the rapid inactivation kinetics of the endogenous conventional PKC isozymes that is driven by rapid metabolism of DG^21^. The high basal activity of PKCη WT and the AD-associated variants was further increased upon UTP treatment and was sustained, consistent with our previous report of novel PKC isozymes’ activity remaining sustained over time after UTP stimulus, driven by persistent DG^21^. The agonist-evoked activity of the variants was not significantly different from PKCη WT (Figure 3d). Taken together, our data suggest that these AD-associated mutations in PKCη exhibit similar activation kinetics as the WT protein but have a lower signaling output assessed with the cytosolic biosensor.

### Enhanced Golgi-associated activity of PKC**η** K65R

Targeting of CKAR to specific intracellular locations has previously established that conventional PKC isozymes signal primarily at the plasma membrane, whereas PKC activity at the Golgi is mainly driven by the novel PKC isozymes^21^. Therefore, we reasoned that the reduced activity of the PKCη variants measured using cytosolic CKAR could reflect enhanced activity at the Golgi which would be sensed with Golgi-targeted CKAR. To test this, we co-expressed Golgi-targeted CKAR and mCherry-PKCη WT or each of the five variants in COS7 cells. Addition of Gö6983 revealed no significant difference in the basal activity of the variants compared with PKCη WT (Figures 4a and 4b). In cells expressing only the reporter, Gö6983 caused a reduction in FRET ratio, reflecting the activity of endogenous novel PKC isozymes in COS7 cells (Figure 4a, grey). However, addition of Gö6983 following UTP stimulation revealed significantly higher PKC activity of the K65R mutant compared with WT (Figure 4c and 4d). Note that in these experiments activity was normalized to the baseline following PKC inhibition. In cells first treated with UTP, the inhibitor-induced reduction in FRET was greater than without UTP treatment because of activation-induced translocation of PKCη to the Golgi. These data show that the PKCη mutation K65R, identified as the most relevant *PRKCH* missense mutation in the AD GWAS, possesses enhanced agonist-induced signaling at the Golgi.

**Figure 4:**
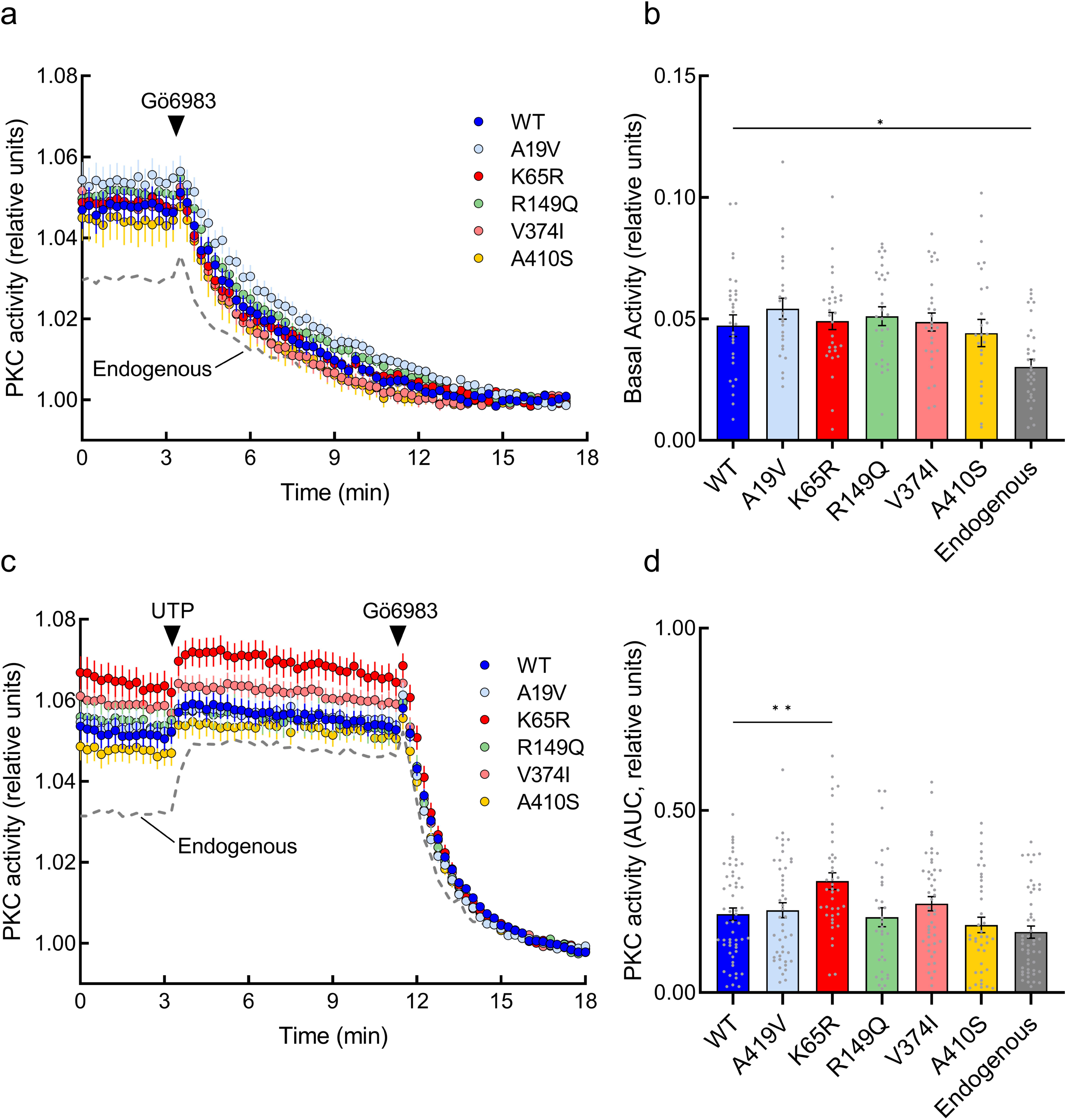
PKCη K65R has enhanced activity at Golgi. **(a)** Normalized FRET ratios representing PKC activity in COS7 cells co-expressing the indicated mCherry-tagged PKCη, or mCherry empty vector (Endogenous PKCs, grey trace) and the CKAR targeted to Golgi. After 3 minutes, cells were treated with 1 μM Gö6983 to study PKC basal activity in the organelle. Data are representative of 26-30 cells per condition, from three independent experiments. **(b)** Relative basal activity quantified using the traces obtained in A by subtraction of the average of the last 2.5 minutes from the average of the baseline registered pre-inhibitor treatment. Data are the mean ± SEM of three independent experiments, *p ≤ 0.05. **(c)** Normalized FRET ratios of PKC activity in COS7 cells co-expressing the indicated mCherry-tagged PKCη, or mCherry empty vector and the Golgi-CKAR. After 3 minutes, PKCs were activated with 100 μM UTP, and subsequently inhibited with 1μM Gö6983. Data are representative of 40-62 cells per condition, from four independent experiments. **(d)** Relative PKC activity, quantified as area under the curve (AUC) from 3 min to 11 min as a measure of signaling output. Data are the mean ± SEM of four independent experiments, **p ≤ 0.001, *p ≤ 0.05.

### The K65R mutation does not alter the intrinsic catalytic activity or substrate binding of PKCη

We next examined the biochemical properties of the PKCη K65R variant to ascertain whether increased catalytic activity could account for its increased signaling output at Golgi. GST-tagged PKCη WT or K65R were purified to homogeneity from insect cells using a baculovirus expression system (Figure 5a). Western blot analysis revealed that both the WT and K65R proteins were processed by the priming phosphorylations at the activation loop (Thr513), the turn motif (Thr656), and the hydrophobic motif (Ser675)^22^ (Figure 5a). Kinase assays revealed that the K65R mutant displayed indistinguishable activation kinetics from PKCη WT when assayed as a function of its two lipid activators DG (half-maximal activation 0.7 ± 0.1 mol% DG vs 0.5 ± 0.1 mol % DG for WT compared with mutant) or phosphatidylserine (PS; half-maximal activation at 3.3 ± 0.1 mol% PS vs 3.7 ± 0.2 mol% PS for WT compared with mutant), or as a function of peptide substrate (K_m_ of 18 ± 3 µM peptide vs 16 ± 3 µM peptide for WT vs mutant) (Figure 5b). These data reveal that the K65R variant has the same degree of autoinhibition (activity was the same in the absence of DG), affinity for ligand (half-maximal activation achieved at the same mol fraction DG), affinity for anionic membranes (half-maximal activation achieved at the same mole % PS), and Km for substrate as PKCη WT. These data establish that the mutation K65R does not alter the intrinsic biochemical properties of PKCη. This is consistent with the residue being surface-exposed and not involved in substrate binding or interdomain contacts that regulate the autoinhibition of the protein.

**Figure 5:**
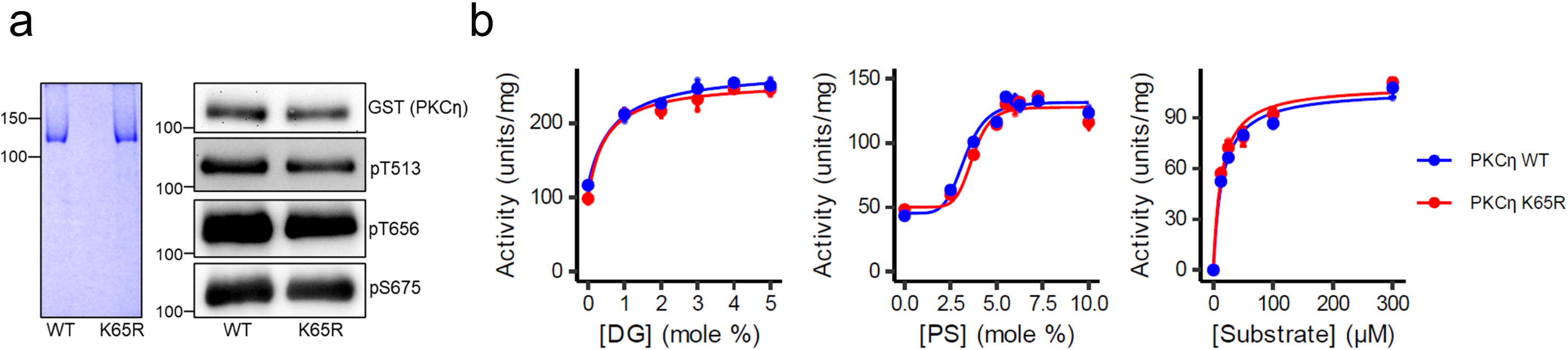
K65R mutation does not alter the intrinsic catalytic properties of PKCη. **(a)** Left: Coomassie Blue-stained SDS/PAGE gel of purified GST-PKCη wild-type and GST-PKCη K65R. Right: Western blot of pure proteins probed with specific antibodies for total PKCη, or the constitutive phosphorylation sites: the activation loop Thr513 (pT513), the turn motif Thr656 (pT656) and the hydrophobic motif Ser675 (pS675)**. (b)** The activity of PKCη WT (blue) or K65R (red) (typically 2.4 nM) was measured as a function of mol% diacylglycerol (DG) or phosphatidylserine (PS), or concentration of peptide substrate, as described in Methods. Data are graphed in units (nmol phosphate per minute) per mg GST-PKC. Data represent the mean ± SD of triplicate samples. Curves represent best nonlinear least squares fit as described in the Methods.

### Enhanced Golgi localization of PKC**η** K65R compared to WT protein

Given that the AD-variants are predicted to be on surface-exposed residues, we reasoned that altered interactions with binding partners could enhance localization of these enzymes to Golgi. To assess this, we co-overexpressed mCherry-tagged PKCη WT or the AD-mutants with the Golgi marker β-1,4-Galactosyltransferase fused to mEGFP (Golgi-mEGFP) and examined their subcellular distribution by confocal microscopy (Figure 6a). Colocalization analysis by Manders’ coefficient^23^, which measures the percentage of mCherry-labeled PKCη that colocalizes with the Golgi marker, revealed significantly increased Golgi localization of the K65R PKCη variant compared with PKCη WT (Figure 6b). A trend towards increased Golgi localization was observed for each variant (Figure 6b), but was only significant (p=0.002) for the K65R. The increased localization of this AD variant with Golgi provides an explanation for its enhanced signaling at this organelle following agonist stimulation. It is noteworthy that the Golgi-localized CKAR did not reveal differences in basal activity of the AD variants. Whether the enhanced PKCη activity of the variants at Golgi is increasing the phosphatase output of cells remains to be determined. Importantly, localization data suggest greater localization at Golgi compared to cytosol for the AD variants compared to WT PKCη.

**Figure 6:**
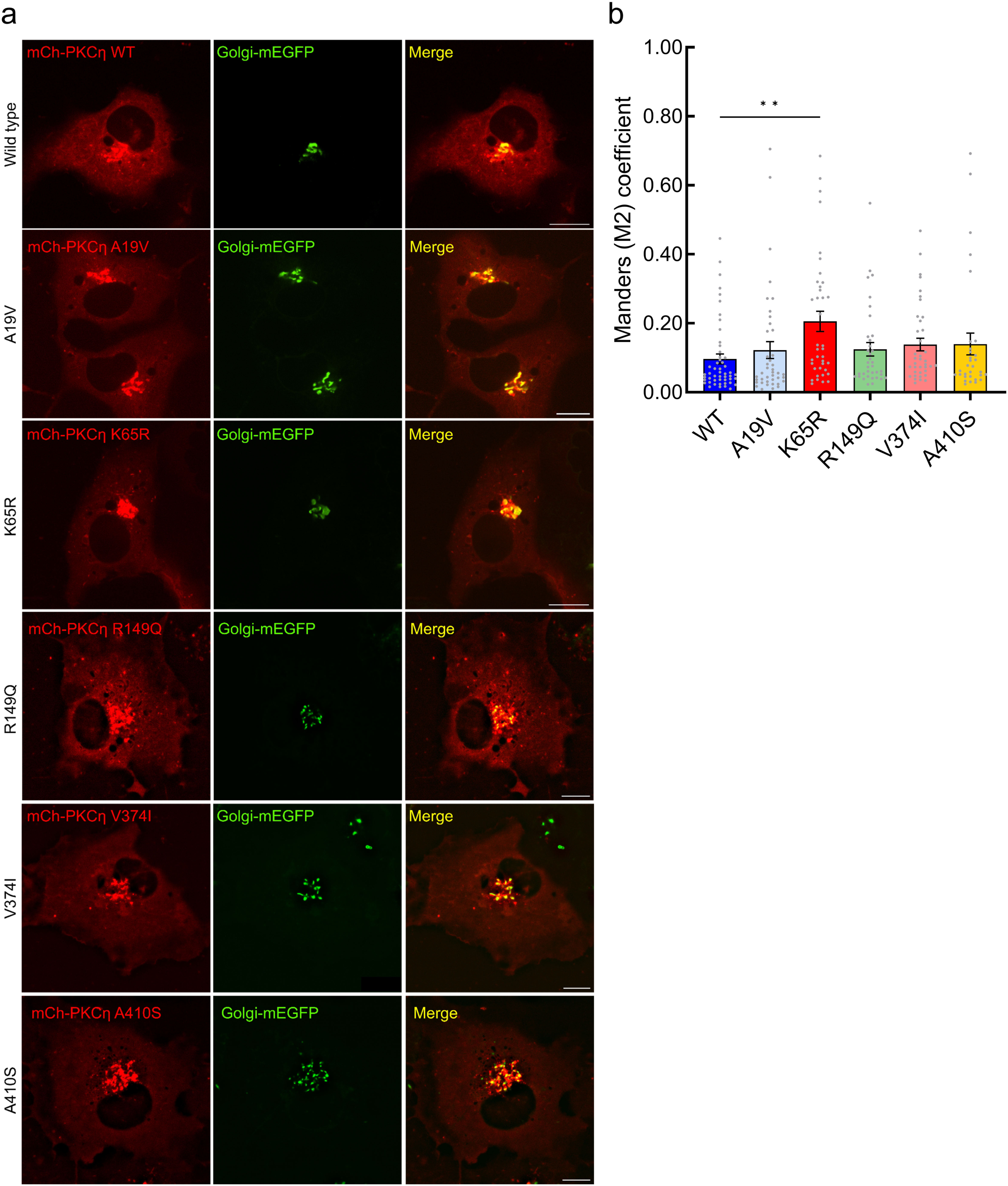
Increased Golgi localization of PKCη K65R compared with WT protein. **(a)** Representative images from confocal microscopy of unstimulated COS7 cells expressing Golgi-mEGFP (green) together with mCherry-PKCη WT or the indicated AD-variant (red). Bar = 10 μm. Images are representative of three independent experiments. **(b)** Bar graph indicating the M2 Mander’s colocalization coefficients^23^ for each mutant or WT PKCη and Golgi marker. Data are the mean ± SEM of three independent experiments,**p = 0.002.

We next assessed whether the increased agonist-evoked activity of PKCη K65R reflected increased agonist-stimulated translocation to the Golgi. We measured the translocation kinetics of YFP-tagged PKCη WT or the AD-variants toward Golgi-tethered CFP (CFP-Golgi) by monitoring the increase in FRET following UTP treatment of cells. Agonist treatment resulted in the greatest translocation to Golgi of the K65R variant, with intermediate levels of translocation of A19V, R149Q, and A410S, and the lowest translocation of WT and V374I (Figure 7). Taken together, our results are consistent with the AD variants having enhanced affinity for Golgi under basal conditions and enhanced translocation following stimulation, an effect most significant and pronounced for the PKCη K65R variant.

**Figure 7:**
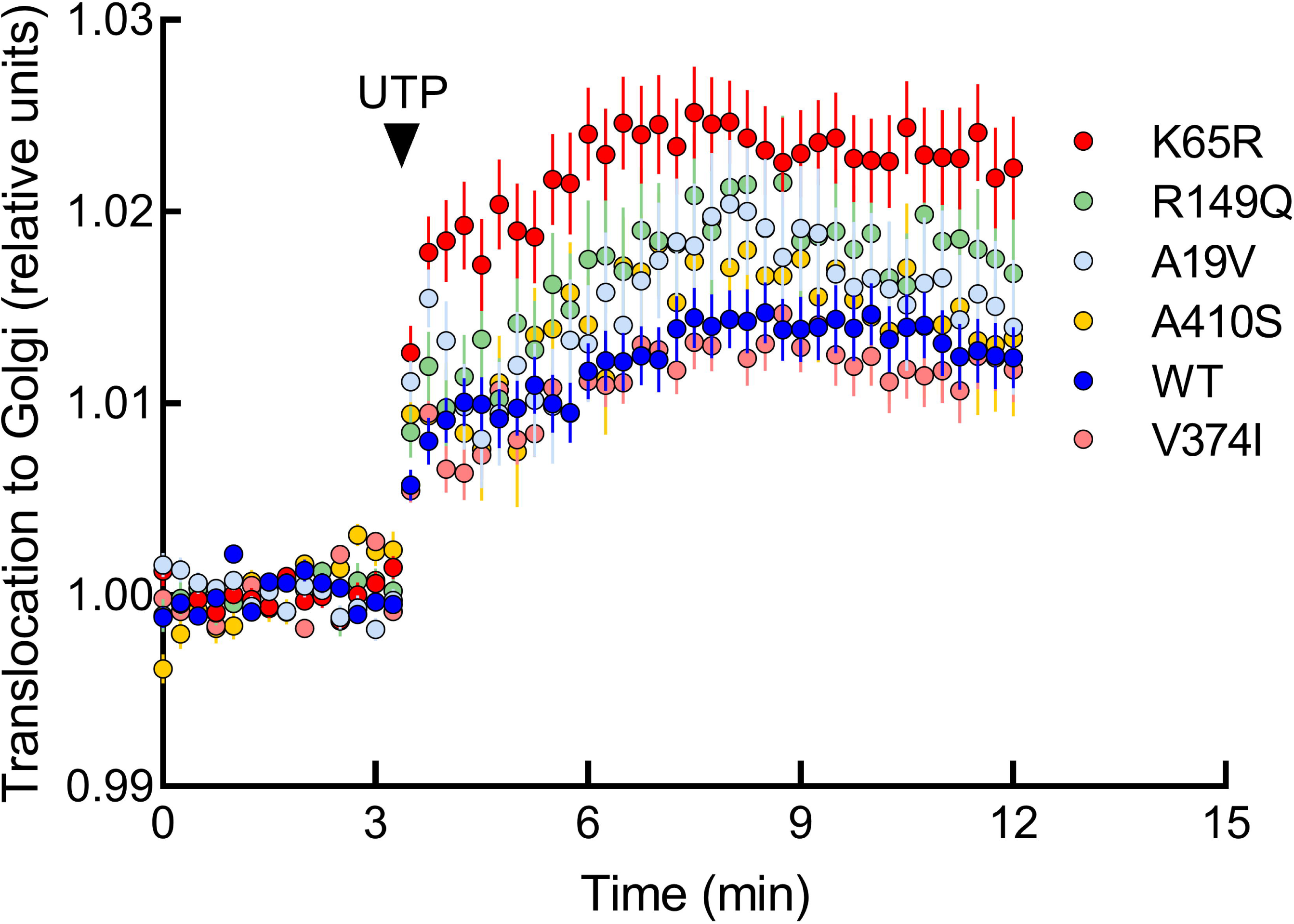
PKCη K65R migrates more to the Golgi after UTP treatment. Traces indicate PKCη translocation to Golgi in COS7 cells co-transfected with Golgi-CFP and the indicated YFP-PKCη. Translocation to plasma membrane was monitored by measuring FRET/CFP ratio changes after stimulation with 100 μM UTP. Data for each cell were normalized to the average of the FRET before UTP addition and represent 27-55 cells per condition. Data are the mean ± SEM of three independent experiments.

### K65R mutation alters interactome of PKC**η**

Given the enhanced Golgi localization of the K65R variant of PKCη, we reasoned that the interactome of PKCη-AD-associated variant would differ from the WT protein one. We performed a proximity-dependent biotin identification (miniTurbo-ID) screen to compare the interactome of WT versus K65R in HEK cells, either before or after UTP treatment^24^. Over 350 protein interactors were identified for PKCη that had differences in abundance between K65R and WT protein. Gene ontology analysis revealed that Golgi-related processes were in the top 10 most significant biological processes based on the proteins, for which binding to PKCη K65R differed from that of WT (Figure 8a). Abundant Golgi-localized proteins were associated with the top 15 most significant biological processes, including signaling enzymes such as protein kinase D (PKD), structural proteins such as the Golgi matrix proteins GOLGA5 and GOLGA4, proteins involved in vesicle-trafficking proteins such as Sec22B and Sec24A, and Rab GTPases such as Rab29. These are primarily proteins associated with the cytoplasmic surface of the Golgi, where PKCη binds. Gene ontology analysis based on mammalian phenotypes identified neurodegeneration as one of the top 10 most significant ontologies, with altered binding detected for proteins such as MARCKSL1 (Figure 8d). Addition of UTP altered the interactome of both the WT and K65R PKCη, with differences between the effect on WT versus the AD variant. Notably, we identified groups of proteins that 1] only bound K65R upon UTP stimulation (Figure 8e, dark grey), 2] only bound WT upon UTP stimulation (Figure 8e, medium grey), 3] bound WT less but K65R better upon UTP stimulation (Figure 8e, light grey), and 4] bound WT more and K65R less upon UTP stimulation (Figure 8e, white). These findings underscore the alteration in surface properties of the K65R mutation having a profound impact on the interactome of the AD variant compared with WT enzyme. Most significantly, they point to a large number of Golgi proteins and processes that are affected by the mutation.

**Figure 8:**
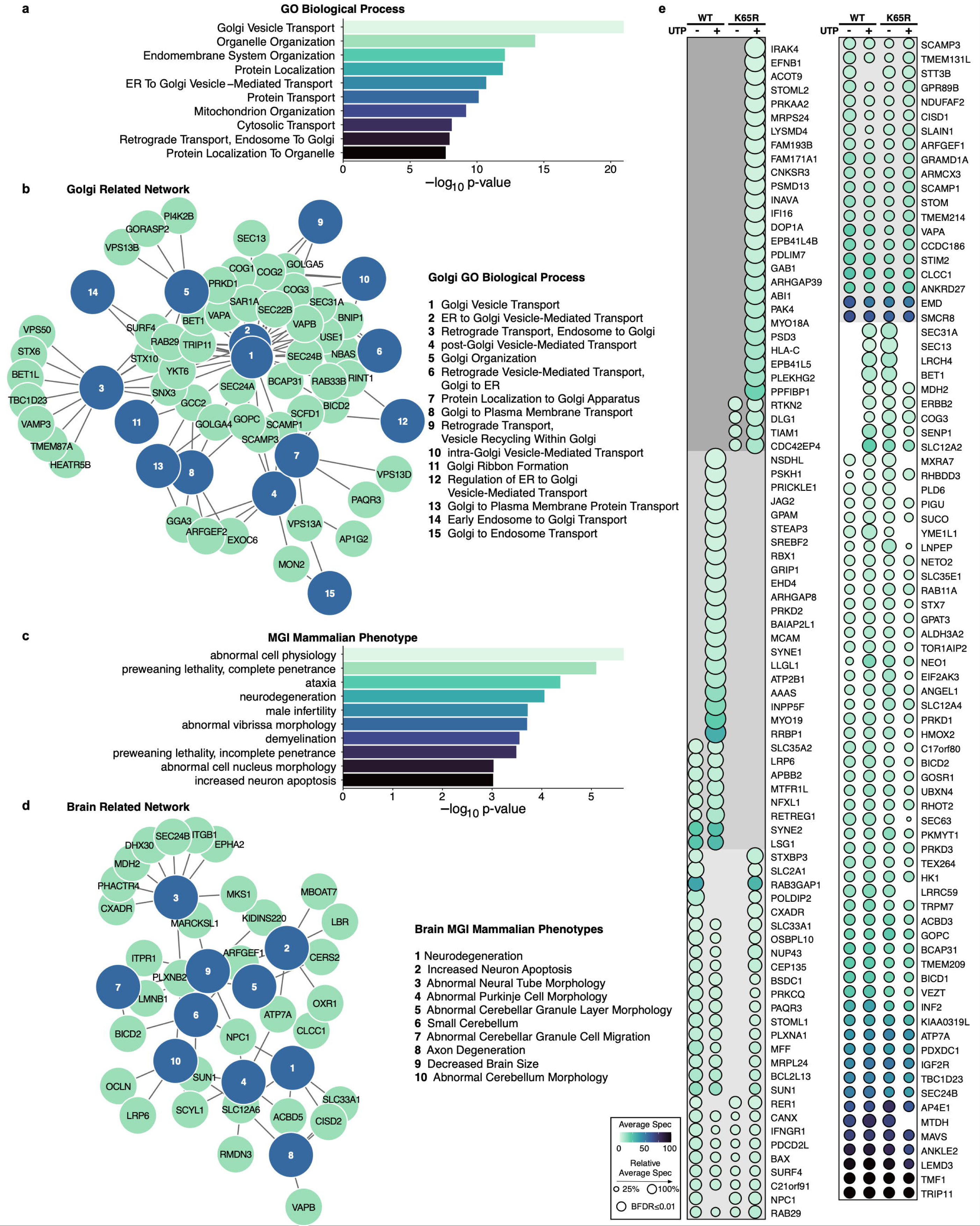
Identification of PKCη WT and K65R protein interactors with miniTurboID. Gene ontology (GO) enrichment analysis identified **(a)** the top 10 most significant ontologies and **(b)** 15 significant Golgi-related processes from GO Biological Process (p ≤ 0.05), and **(c)** the top 10 most significant ontologies and **(d)** 10 significant brain-related phenotypes from MGI Mammalian Phenotypes (p ≤ 0.05) for protein interactors altered between PKCη WT and K65R (BFDR ≤ 0.01). **(e)** Dot plot displays all protein interactors that have opposite changes in abundance with UTP compared to vehicle treatment between PKCη WT and K65R (different directionality is designated by grey shading). Colormap indicates abundance (average spectral count; spec), dot size indicates relative abundance, and outline color indicates BFDR ≤ 0.01.

### *PRKCH* expression is increased in the brain of AD patients

Lastly, analysis of the GTEx database revealed increased expression of *PRKCH* in several brain tissues (most pronounced in amygdala) for the recessive allele of rs7161410 (AA) compared to GA and GG alleles (1.087 vs. -0.07720, -0.03857, respectively, Supplementary Figure 2). Assuming mRNA correlates with protein expression, this would support gain of function of K65R in AD, with our functional data indicating aberrantly high signaling at the Golgi.

## Discussion

We identified an intronic variant in the *PRKCH* gene strongly associated with AD risk and in strong LD with a rare missense mutation that alters PKCη function, by strategically using a family-based GWAS under a RM using WGS from AD affected families. Live-cell analysis revealed that several missense mutations in linkage disequilibrium with the AD-associated *PRKCH* intronic variant risk allele exhibited reduced cellular basal activity in the cytosol; however, the genetically most relevant mutation, K65R, showed enhanced localization and signaling at the Golgi, as compared to WT PKCη. Biochemical analysis of K65R revealed that the mutation did not alter its intrinsic biochemical properties. Rather, this mutation enhanced association of PKCη with Golgi and altered its interactome. These findings illustrate the power of family-based GWAS of WGS data using a RM, to identify *bona fide* functional targets for AD.

By design our GWAS could eliminate false-positive findings in terms of quality control and is robust to population stratification. In our analyses, the variants rs4420638 and rs56131196 were suggested as the most significant loci which confirms previously well-known susceptibility region for AD on chromosome 19q13 (i.e., *APOE* cluster gene region) ^25,26^. Along with *APOE*, our analysis showed that the association of *PRKCH* with AD affected status was much stronger under a recessive versus additive inheritance model (Figures 1c and 1d). Previously, we reported association of AD with rare-variant signals in *PRKCH* using single-variant and spatial-clustering analyses^27^.

PKCη is primarily expressed in microglia as evidenced by the brain cell type-specific enhancer-promotor interactome map of active promoters^28^. Microglia are a macrophage population in the brain that serve as the primary immune effector cells^29^, and their dysregulation is associated with the pathogenesis of neurodegenerative diseases such as AD^30,31^. In addition, dozens of AD-associated genes are known to impact microglial function in the brain^32^. Notably, the phospholipase C γ gene, *PLCG2*, which produces DG to activate PKC, is a known AD risk gene that is highly expressed in microglia^33^. Similarly, the AD risk factor TREM2 has been proposed to modulate the inflammatory signaling mediated by PKC^33,34^.

The high expression of PKCη in microglia, coupled with its known role in immune and inflammatory signaling, poise this PKC isozyme in a prime position to regulate microglial-mediated inflammation, a key contributor to the progression of AD^35,36^. Furthermore, a germline mutation V374I which increases the activity of PKCη is associated with stroke: the SNP, rs2230500 is strongly associated with the risk of ischemic stroke or cerebral hemorrhage in the Asian population ^37–39^, which has a *p*-value of 0.011 (0.033 in NIMH only) with the same direction as rs7161410 in our additive model analysis, despite the small affected family size (Figure 2a). The correlation between stroke and AD^40^ provides another link between deregulated PKCη function and AD. One plausible mechanism based on coronary arterial specimens suggested that PKCη is expressed in not only vascular endothelial cells but also white blood cells, of which foamy macrophages contribute to the cellular uptake of lipoproteins, mainly low-density lipoproteins (LDL)^37,41^. Amyloid-β also enhances cellular cholesterol accumulation by opsonization of LDL which accelerates macrophage foam cell formation^41^. Therefore, PKCη signaling may be implicated in the potential participation of cerebral perivascular macrophages and microglia leading to the disturbance of the cholesterol balance of brain tissues, thereby promoting amyloid accumulation and plaque generation. Lastly, a recent study revealed that PKCη is highly enriched in cortical astrocytes, near amyloid plaques in the 5XFAD mouse model of AD, proposing that PKCη regulates neuroinflammation in AD^42^. Thus, considerable evidence supports the possibility that restoring normal function of PKCη is a potential therapeutic target in AD.

This study is not without limitations. First, we acknowledge that the top recessive SNP, rs7161410, reaches only suggestive genome-wide significance (p<=5×10^-7^) in our discovery dataset. We have made several unsuccessful attempts to replicate this recessive signal in multiple other unrelated case-control cohorts and large biobanks. We note possible phenotype differences between family-based AD, sporadic AD in case-control cohorts and AD-by-proxy or ICD10 in large biobanks and the limited availability of other family-based cohorts with multiplex AD families. Second, the rare functional mutation K65R, carriers of which were homozygous for rs7161410, was present in only one informative non-Hispanic white family, contributing to the rs7161410 signal. However, we observed nominal additive association for the functional mutation K65R (p_add_=0.030) in the latest NIA ADSP case-control dataset. In addition, we have shown experimentally how this functional mutation leads to enhanced localization and signaling of the protein at Golgi. Given that the recessive rs7161410 signal was most pronounced in a subset of Dominican families, further functional research and replication attempts are necessary specifically in Dominican subjects. Next, our biochemical and cellular studies in COS and HEK cells, revealed K65R mutation in PKCη enhanced signaling at the Golgi, and altered interaction with Golgi-associated proteins. These data are consistent with a long history of reports on dysregulated Golgi structure and function in AD^44,45^. It is noteworthy that one of the proteins that displayed enhanced binding to K65R compared to WT was Rab29, which is associated with Parkinson’s Disease by recruiting the LRRK2 kinase^46^. Future studies examining how PKCη regulates Golgi function, particularly in microglial cells, would be important to better understand how PKCη is a risk factor in AD. Additionally, whether the intronic variant that first drew our attention to PKCη results in altered protein expression will be important to determine in order to understand how it confers a risk factor for AD.

In summary, the association of an intronic *PRKCH* variant with increased AD risk using a RM in a family-based GWAS of WGS data, along with a missense mutation in strong LD with the *PRKCH* risk allele that alters PKCη signaling at the Golgi, suggests a novel biological link of AD etiology and neuropathogenesis with PKCη. Our results also support previous studies demonstrating a role for altered PKC signal activity in AD pathogenesis. Future studies aimed at understanding the mechanisms underlying abnormal localization and signaling of PKCη at the Golgi will be needed to help inform therapeutic strategies aimed at preventing and treating AD.

## Materials and Methods

### Discovery whole genome sequencing analysis

The samples from the NIMH AD Initiative were sequenced on the Illumina HiSeq 2000 platform and aligned to the human reference genome (GRCh38). Further details are discussed in ^47^. Sequencing data for the NIA cohort was obtained from the National Institute on Aging Genetics of Alzheimer’s Disease Data Storage Site (NIAGADS) under accession number: NG00067.v9. A subject was considered to be affected if he/she was included in one of the following categories: “Definite AD”, “Probable AD” or “Possible AD”. Unaffected subjects had either no dementia, suspected dementia (34 subjects) or non-AD dementia (4 subjects). It is important to note that NIA ADSP families by design did not include individuals with two APOE-ε4 alleles. After standard quality control, both cohorts were merged together. For variant quality control, we excluded multi-allelic variants, monomorphic variants, singletons (i.e. variants with only one alternative allele across the dataset), indels, and variants which had one missing allele among 2 alleles in an individual. The remaining variants were filtered based on Mendel errors, genotyping rate (95%) and Hardy-Weinberg equilibrium (p > 10^−6^). GWAS was conducted using the FBAT software (version 2.04) under RM and AM^48^. We set the threshold for “suggestive” genome-wide statistical significance at p < 5 x 10^−7^. Although FBAT is robust to population structure and phenotype misspecification, we have performed a stratified analysis by population. There were 412 subjects of Dominican ancestry, 122 subjects from a Dutch isolate group,18 subjects of Puerto Rican ancestry, 84 subjects of African-American ancestry and the rest was of nonHispanic white ancestry. For replication, we used unrelated case-control subjects from three datasets: ADSP unrelated case-control subjects (clinically defined AD phenotype), UK Biobank (AD-by-proxy phenotype) and AllofUS (AD-by-proxy phenotype). Interaction analysis was performed using the “fbati” R package with the “fbatge” function.

### Replication in ADSP unrelated case-control dataset

We have obtained whole genome sequencing data for unrelated case-controls from the NIAGADS webpage (ADSP R4). After sample quality control the full dataset contained 25,660 subjects (ncases=10,565). Subpopulations were defined using self-reporting and principal components which were calculated based on rare variants using the Jaccard index^49^. Outliers more than 5 standard deviations away based on 10 principal components were excluded. In each subpopulation (European Americans, N=9,609, N_cases_=6,136; African Americans, N=4,263, N_cases_=1,405; Hispanic subpopulation, N=8,466, N_cases_=2,559; Asian subpopulation, N=2,543, Ncases=181), we performed a logistic regression using the RM or AM of inheritance for case/control status as implemented in PLINK 2^50^. We included sex, age, sequencing center, and 5 Jaccard principal components with standardized variance as covariates.

### Replication in UK Biobank and AllofUS biobank

We have used whole genome sequencing data from 200,000 subjects available in UK Biobank ^51^ and from 245,388 subjects in AllofUS ^52^. AD-by-proxy phenotype was defined as having family history of AD (at least one affected parent). Subjects with unavailable information on family history were excluded. We performed a whole genome sequencing regression with covariates as implemented in regenie ^53^. Step 1 was performed on SNPs from array data and we followed the regenie recommendations (https://rgcgithub.github.io/regenie/recommendations). In step 2 we included age at enrollment, sex, twenty principal components (PCs) and sequencing center (available only in UK Biobank). PLINK2 was used to extract the SNPs of interest. PCs for UKB were provided. PCs for AoU were obtained from a LD-pruned subset of SNPs using array data.

### Plasmids and Reagents

The CKAR2^20^, Golgi-CKAR and Golgi-CFP^21^ plasmids were described previously. Human PKCη was YFP-, HA- or mCherry-tagged at the N-terminus in pcDNA3 vectors using Gateway Cloning (Life Technologies). All AD-associated mutants were generated using QuickChange site-directed mutagenesis (Agilent) following the manufacturer’s instructions. Golgi-mEGFP was obtained from AddGene (ID no. 182877). Uridine-5 -triphosphate (UTP) Trisodium Salt was obtained from MerckMillipore (cat no. 6701). Gö6983 (catalog no. 285) was purchased from Tocris. The anti-phosphorylated PKCη (pT655) antibody was purchased from Abcam (Ab5798) and pan anti-phosphorylated PKC hydrophobic motif (βII pS660) antibody was from Cell Signaling Technology (9371S). The pan anti-phosphorylated-PKC activation loop antibody was previously described ^54^. The total PKCη (C-15, sc-215) and GST (B-14, sc-138) antibodies were purchased from Santa Cruz Biotechnology. HRP-conjugated anti-rabbit (catalog no. 401315) and anti-mouse (catalog no. 401215) secondary antibodies and BSA (catalog no. 12659) were from Millipore. All antibodies were diluted in 1% BSA dissolved in PBS-T (1.5 mM Sodium Phosphate Monobasic, 8 mM Sodium Phosphate Dibasic, 150 mM NaCl, 0.05% Tween-20) with 0.25 mM thimerosal (Thermo Scientific, catalog no. J61799.14). Bradford reagent (catalog no. 500-0006), protein standards ladder (catalog no. 161-0394), bis/acrylamide solution (catalog no. 161-0156), and polyvinylidene difluoride (PVDF) (catalog no. 162-0177) were purchased from Bio-Rad. Luminol (catalog no. A-8511) and p-coumaric acid (catalog no. C-9008) used to make chemiluminescent substrate solution were purchased from Sigma-Aldrich. Lipids used in kinase assays (DG, 800811C and PS, 840034C) were from Avanti Polar Lipids.

### Cell Culture and Transfection

COS7 cells were maintained in Dulbecco’s modified Eagle’s medium (Corning, catalog no. 10-013-CV) containing 10% fetal bovine serum (Atlanta Biologicals, catalog no. S11150) and 1% penicillin/streptomycin (Gibco, catalog no. 15-140-122) at 37°C in 5% CO_2_. Cells were periodically tested for Mycoplasma contamination by a PCR-based method. Transient transfections were carried out using a Lipofectamine 3000 kit (Thermo Fisher Scientific) per the manufacturer’s instructions, and constructs were allowed to express for 24 hours prior to imaging experiments.

### FRET Imaging and Analysis

COS7 cells were seeded were seeded into plates (Corning, catalog no. 430165) containing glass cover slips (Fisherbrand, catalog no. 12545102) glued on using SYLGARD 184 Silicone Elastomer Kit (Dow, catalog no. 04019862). For CKAR assays, cells were co-transfected with 1 μg mCherry-PKCη wild type or each one of the PKCη AD-mutants constructs, together with 1 μg CKAR2 or Golgi-CKAR DNA. For translocation assays, cells were co-transfected with 500 ng YFP-tagged PKCη and 500 ng Golgi-CFP. 24 hours post-transfection, cells were imaged in 2 mL Hank’s Balanced Salt Solution (Corning, catalog no. 21-022-CV) with 1 mM CaCl_2_ added fresh prior to imaging. Images were acquired on a Zeiss Axiovert 200M microscope (Carl Zeiss Micro-Imaging Inc.) using an Andor iXonUltra 888 digital camera (Oxford Instruments) controlled by MetaFluor software (Molecular Devices) version 7.10.1.161. Images were acquired every 15 seconds, and baseline images were acquired for 3 minutes. Drugs were added dropwise to the dish in-between acquisitions. For CKAR (cytosolic and Golgi) activity assays, cells with equal mCherry and YFP levels were selected for analysis and the FRET ratios for each cell were normalized to the average of the last 10 cycles. Basal activity was calculated as the subtraction of the average of the last 10 cycles from the first 3 minutes prior to drug addition. For translocation assays, FRET ratios for each cell were normalized to the average of the first 3 minutes prior to drug treatment.

### Confocal Microscopy of Live Cells

COS7 cells were seeded onto Mat Tek glass bottom culture dishes (catalog no. P35G-1.0-14-C), the next day cells were transfected with 1 μg of Golgi-mEGPF together with mCherry-PKCη WT or the AD-mutants. After 24 h the live cells were imaged by confocal microscopy. Spinning disk confocal microscope specifications: Yokogawa X1 confocal scanhead mounted to a Nikon Ti2 microscope with a Plan apo lambda 100x oil NA 1.45 objective, microscope controlled via NIS Elements using the 405 nm, 488 nm, and 640 nm lines of a four-line (405 nm, 488 nm, 561 nm, and 640 nm) LUN-F-XL laser engine and a Prime95B camera (Photometrics). Image channels were acquired using bandpass filters for each channel (455/50, 525/50 and 705/72). Images were analyzed using NIS Elements software (Nikon) and JACoP plugin^55^ in ImageJ (NIH) was utilized to perform the colocalization analysis.

### Quantification and Statistical Analysis

For imaging experiments, intensity values and FRET ratios were acquired using MetaFluor software and normalized as described above. Statistical tests were performed using Prism (GraphPad Software) version 9.5.0. Structures were modeled using PyMOL version 2.3.0 (Schrödinger, LLC).

### Insect Cell Culture, Protein Purification, and Kinase Assays

Sf-9 insect cells were employed to obtain baculoviruses expressing human GST-PKCη using the Bac-to-Bac expression system (Invitrogen). Human GST-PKCη WT and K65R proteins were expressed and purified with glutathione Sepharose beads as previously described ^43,56^, with the following modifications. Cells were centrifuged, washed, and lysed in 50 mM Hepes, pH 7.5, 1 mM EDTA, 100 mM NaCl, 1% Triton X-100, 100 µM PMSF, 1 mM DTT, 2 mM benzamidine, 50 µg/mL leupeptin, and 1 µM microcystin. The soluble lysate was incubated with glutathione resin beads (EMD Millipore, #70541-4) for 1 hour at 4°C on a nutator. Protein-bound beads were washed three times in wash buffer (50 mM Hepes, pH 7.5, 1 mM EDTA, 100 mM NaCl, 1 mM DTT) and eluted five times in wash buffer with 10 mM glutathione. The purified protein was concentrated in 50 kDa Amicon centrifugal filter unit (EMD Millipore, #905024) and exchanged into 20 mM Hepes, pH 7.5, 1 mM EDTA, 1 mM EGTA, and 1 mM DTT. Glycerol was added to a final concentration of 50% for the proper storage of the enzyme at −20 °C.

The activity of purified GST-PKCη (2.6 nM) was assayed toward a peptide substrate based on the predicted pseudosubstrate site of PKCη with a Ser in substitution of an Ala (Ac-RKRQRSMRRRVH-NH2, from GenScript). The kinase assay was performed as described previously^43^ with the following changes in the protocol: the standard conditions of the assay were: 100 µM ATP, 10 µCi/ml [γ-32P] ATP, 100 µM substrate, 5 mM MgCl_2_, 500 µM EGTA, 0.06 mg/mL BSA, and Triton X-100 (0.1% wt/vol) mixed micelles containing 15 mol % PS and 5 mol % DG in 50 mM Hepes, pH 7.5, and 1 mM DTT. Mol% PS, mol% DG and substrate concentration were adjusted as described in figure legend. One unit of activity is defined as 1 nmol phosphate incorporated per min into substrate. Kinase assay data were fit in R (version 4.4.2, 2024-10-31) using nonlinear least squares regression using the nls() command in the base package *stats*. Dose-response curves were fit to a modified Hill equation of the form “activity = a0 + (agonist^n) * (a1 - a0)/(agonist^n + EC50^n)” where a0 equals minimal (unstimulated) activity, a1 equals maximal activity, agonist equals agonist concentration in mole percent, EC50 equals the agonist concentration at which half-maximal induced activity is realized, and n is equivalent to the Hill coefficient^57^. For diacylglycerol curves, n was constrained to 1 to indicate the non-cooperativity of this binding event. For phosphatidylserine curves, n was estimated from the data. Substrate curves were fit to the Michaelis-Menten equation of the form “activity = (Vmax*substrate)/(Km + substrate)” where Vmax indicates the maximal activity, substrate indicates the substrate concentration in micromolar, and Km is the concentration of substrate at which the reaction rate is half of Vmax. Data wer visualized using the R-packages *ggplot* (version 3.5.1) and *ggpubr* (version 0.6.0).

### eQTL analysis

We also explored the association between the variant and potential candidate genes by cis-expression quantitative trait loci (eQTL) in brain tissues based on the GTEx release version 8 database (https://www.gtexportal.org/home). The R statistical software (http://www.R-project.org) was used to evaluate these tests.

### MiniTurboID

#### Cell lines generation

pENTR-PKCη-WT or pENTR-PKCη-K65R constructs were fused to pDEST-pcDNA5-miniTurboID-3xFLAG-N-term via Gateway cloning (Thermo Fischer Scientific) following the manufacturer’s specifications. These constructs were used to generate stable cell lines in HeLa Flp-In T-REx cell pools, as described in ^58^.

#### Expression induction, drug treatment and cell collection

Stable cell lines (miniTurboID-PKCηWT, miniTurboID-PKCηK65R, miniTurboID empty vector and parental cell line) were seeded in 15 cm dishes. When cells reached 80% confluency were treated with 1 mg/ml Doxycycline for 24h to induce expression. All plates were then treated for 1 h with 50 mM Biotin, half of them were treated with 100 mM UTP and the other half with ddH_2_O as vehicle control. Cells were subsequently washed twice with ice-cold PBS, collected with scrapper into 1.5 mL PBS, and centrifuged at 500 x g for 5 min. Supernatants were carefully removed and cell pellets were stored at -70□C until cell lysis.

#### Cell lysis

Each frozen cell pellet was resuspended in ice-cold modified RIPA lysis buffer (50 mM Tris-HCl, pH 7.4, 150 mM NaCl, 1 mM EGTA, 0.5 mM EDTA, 1 mM MgCl2, 1% NP40, 0.1% SDS, 0.4% sodium deoxycholate, 1 mM PMSF, 1x Protease Inhibitor cocktail) at a 1:4 pellet weight: volume ratio. Cells were sonicated for 15 sec (5 sec on, 3 sec off for three cycles) at 30% amplitude on a sonicator with 1/8” micortip. Samples were kept on ice. 250 U of TurboNuclease and 10 µg of RNase were added, and tubes rotated (end-over-end) at 4°C for 15 min. Subsequently, SDS concentration was increased to 0.4% (by the addition of 10% SDS) and rotated at 4°C for 5 min. Lysates were centrifuged at 20,817x g for 20 min at 4°C and supernatant was transferred to a 2 ml centrifuge tube.

#### Streptavidin purification

A master mix of streptavidin beads was prepared by using 35 μl of slurry (20 µl bed volume) for each sample plus 10% to account for any losses. Streptavidin sepharose beads were washed 3 times with 1 ml lysis buffer (minus proteinase inhibitors, PMSF and deoxycholate). For beads washing, tubed were mixed by inversion, centrifuged 400x g for 1 min and supernatant was discarded. Following the last wash, beads were resuspended as a 50% slurry. 40 μl of the 50% slurry of streptavidin beads was added to the clarified supernatant and rotated using gentle end-over-end rotation for 3 hours at 4 C. Beads were pelleted by centrifugation at 400x g for 1 min at 4 C, supernatant discarded and beads were transferred in 1 ml fresh RIPA-wash buffer (50 mM Tris-HCl, pH 7.4, 150 mM NaCl, 1 mM EDTA, 1% NP40, 0.1% SDS, 0.4% sodium deoxycholate) to a new microcentrifuge tube (this removes ‘sticky’ proteins bound to side of tube). Beads were washed once with SDS-Wash buffer (25 mM Tris-HCl, pH 7.4, 2% SDS), twice with RIPA-wash buffer, once with TNNE buffer (25 mM Tris-HCl, pH 7.4, 150 mM NaCl, 0.1% NP40, 1 mM EDTA) and three times with 50 mM ammonium bicarbonate (ABC) buffer, pH 8.0. For each wash step, tubes were mixed by inversion, centrifuged 400x g for 1 min and supernatant was discarded.

#### Trypsin Digest

Residual ABC buffer was removed by pipette, beads resuspended in 70 µl of 50 mM ABC buffer and 1 µg trypsin dissolved in 50 mM ABC buffer was added and incubated at 37 C overnight with agitation. Additional 0.5 µg of trypsin was then added and incubated for a further 3 hours. Beads were centrifuged (400x g, 2 min) and supernatants collected in a new 1.5 ml tube. Beads were washed twice with 150 µl mass spectrometry grade H_2_0 (pelleting beads in between) and the wash supernatant was pooled with the peptide supernatant. Supernatants were centrifuged at 16,100x g for 10 min and most of the supernatant transferred (leaving ∼30 µl residual so as not to transfer beads) to a new tube. Pooled supernatant was lyophilized using vacuum centrifugation without heat. Stored dried peptides at -40 C until ready for mass spectrometry analysis.

#### Mass spectrometry analysis

(i) Instrument: Bruker timsTOF Pro 2. (ii) Method: timsTOF dda-PASEF (22min gradient-60SPD): For data-dependent acquisition (DDA) LC-MS/MS, One-sixteenth of digested peptides were analyzed using a nano-HPLC (High-performance liquid chromatography) coupled to MS. One-sixteenth of the sample was loaded onto Evotip Pure per manufacturer instructions. Peptides were eluted from the Performance column (cat#: EV-1109, 8cmx150µm with 1.5µm beads), heated at 40 C) from a 20µm diameter emitter tip with the 60SPD pre-formed acetonitrile gradient generated by an Evosep One system, and analyzed on a timsTOF Pro 2. MS1 scans were performed from 100-1700Da in PASEF mode with an accumulation and ramp time of 100ms (with 4 PASEF ramps and active exclusion at 0.4min), and within the mobility range (1/K0) of 0.85 to 1.3V·s/cm2. The total cycle time was of 0.53s. The target intensity was set to 17,500 and intensity threshold set to 1750. 1+ ions are excluded from fragmentation using a polygonal filter. The auto calibration was off.

#### Data analysis

Data files were searched with MSFragger^59^ 3.7 within the ProHits^60^ LIMS using FASTA database from UP000005640 human Uniprot (no isoforms). The data was searched for peptides digested with trypsin, with a maximum of 2 missed cleavages. Acetylated protein N-term and oxidated methionine were set as variable modifications with precursor and fragment mass tolerance set to 40 ppm. MSBooster and Percolator were turned on. Percolator required a minimum probability of 0.5 and did not remove redundant peptides. The target-decoy competition method was used to assign q-values and PEPs. For ProteinProphet, the maximum peptide mass difference was set to 30ppm. When generating the final report, the protein FDR filter was set to 0.01. FDR was estimated by using both filtered PSM and protein lists. Razor peptides were used for protein FDR scoring. All other parameters were default.

#### Data processing and visualization

Significance Analysis of INTeractome (SAINT) software package was used to identify and score protein-protein interactions^61^. SAINT was used to identify high-confidence protein interactors versus control samples. Prior to applying SAINT, proteins were filtered based on unique peptides ≥ 2 to ensure confidence in identified proteins. Identified proteins had a Bayesian False Discovery Rate (BFDR) ≤ 0.01 and are considered high-confidence protein interactors. R Programming was used for subsequent data processing^62^. Protein interactors with altered abundance, measured as change in average spectral count, between genotypes (PKCη WT and PKCη K65R) and treatments (vehicle and UTP) were identified. Gene ontology (GO) enrichment analysis was performed for protein interactors that were altered between PKCη WT and PKCη K65R vehicle treated groups using EnrichR to identify biological features from annotated gene databases, including GO Biological Process 2023 and MGI Mammalian Phenotypes Level 4 2021^63–67^. From the identified ontologies that showed significant enrichment for the protein interactors (p ≤ 0.05), Golgi-related or brain-related ontologies were selected for visualization in a network analysis performed using Igraph^68^. ProHits-viz was used to visualize protein-protein interaction data with dot plots. Representative dot plot displays protein interactors in which abundance that was increased with UTP treatment in PKCη WT and decreased with UTP treatment in PKCη K65R and vice versa ^60,69^. Dots are absent in the plot for proteins that were below detection or failed to pass a BFDR ≤ 0.01.

## Supplementary Materials

Supplementary Table 1. Top SNPs associated with Alzheimer’s disease with p-values stronger than suggestive genome-wide significance (P < 5×10-7) under recessive model and their corresponding outcomes under additive model.

Supplementary Table 2. FBAT results for rs7161410 under recessive model with and without families with K65R mutation carriers.

Supplementary Table 3. Number of carriers of functional variants in *PRKCH* for each genotype of rs7161410.

Supplementary Figure 1: Pedigree of K65R carriers in the discovery family-based dataset.

Supplementary Figure 2: *PRKCH* expression increases in the brains of AD patients.

Supplementary Acknowledgements.

## Acknowledgements

The authors wish to thank Cassandra Wong and Laura McGary at the Network Biology Collaborative Centre Proteomics Facility for the miniTurboID analysis. We thank Dr. Peng Guo and the Nikon Imaging Center at UCSD for the support on microscopy experiments. The computations in this paper were run in part on the FASRC Cannon cluster supported by the FAS Division of Science Research Computing Group at Harvard University. The funding body has no role in the design of the study and collection, analysis, and interpretation of data and in writing the manuscript. Please refer to the Supplementary Note for full acknowledgements.

## Data availability

NIA ADSP WGS dataset is available from DSS NIAGADS under accession number: NG00067. The NIA ADSP dataset contains data in part obtained from the Alzheimer’s Disease Neuroimaging Initiative (ADNI) database (adni.loni.usc.edu). As such, the investigators within the ADNI contributed to the design and implementation of ADNI and/or provided data but did not participate in analysis or writing of this report. The NIMH dataset analyzed during the current study will be deposited to dbGAP upon publication. UKB data access is available through application at https://ukbiobank.dnanexus.com/landing. This research was conducted using the UKB application number 81874. Access to individual-level data from the All of Us research program was obtained through an MGB-signed a data use agreement with All of Us (https://www.researchallofus.org/register/).

## Consent statement

All participants provided electronically signed consent. UK Biobank received ethical approval from the NHS North West Centre for Research Ethics Committee with the latest renewal in 2021 (Ref: 11/NW/0382). Massachusetts General Hospital has a Data Use and Registration Agreement with AllofUS. This study was approved by the relevant Institutional Review Board from Massachusetts General Hospital (protocol number 2022P000614, 2015P000111, 2019P001915).

## Author contributions

Conceptualization: CL, ACN, RET.

Methodology: MCG, DP, SL, SW, JH, GH, DLD, WH, LB, CL, ACN, RET.

Investigation: MCG, DP, SL, SW, JH, JW, MW, GL, JM, YY, IC, SM, GH, DLD, WH, LB, CL, ACN, RET.

Funding acquisition: CL, ACN, RET.

Project administration: KM. Supervision: CL, ACN, RET.

Writing – original draft: MCG, DP, SL, SW, CL, ACN, RET.

Writing – review & editing: MCG, DP, SL, SW, JH, JW, MW, GL, JM, YY, IC, KM, SM, GH, DLD, WH, LB, CL, ACN, RET.

