## Supplementary Material for "Protein kinase C eta enhances Golgi-localized signaling and is associated with Alzheimer’s disease using a recessive mode of inheritance"

**Supplementary Tables**

**Supplementary Table 1.** Top SNPs associated with Alzheimer’s disease with *p*-values stronger than suggestive genome-wide significance (P < 5×10^-7^) under recessive model and their corresponding outcomes under additive model.

| Model | rsID | Chr | Position | Ref | Alt | Gene / Consequence | afreq | fam | S-E(S) | Var(S) | Z | *p*-value |
| --- | --- | --- | --- | --- | --- | --- | --- | --- | --- | --- | --- | --- |
| Recessive model | rs7161410 | 14 | 61325380 | G | A | PRKCH : Intron Variant | 0.261 | 36 | 18.26 | 12.034 | 5.264 | 1.41E-07 |
|  | rs56131196 | 19 | 44919589 | G | A | APOC1 : 500B Downstream Variant | 0.336 | 63 | 27.45 | 17.154 | 6.628 | 3.48E-11 |
|  | rs4420638 | 19 | 44919689 | A | G | APOC1 : 500B Downstream Variant | 0.337 | 63 | 27.45 | 17.154 | 6.628 | 3.48E-11 |
|  | rs12721051 | 19 | 44918903 | C | G | APOC1 : Intron Variant | 0.324 | 61 | 26.617 | 16.682 | 6.517 | 7.33E-11 |
|  | rs429358 | 19 | 44908684 | T | C | APOE : Missense Variant | 0.33 | 54 | 23.15 | 15.495 | 5.881 | 4.18E-09 |
|  | rs10414043 | 19 | 44912456 | G | A | APOC1 : 2KB Upstream Variant | 0.262 | 38 | 18.212 | 10.609 | 5.591 | 2.25E-08 |
|  | rs769449 | 19 | 44906745 | G | A | APOE : Intron Variant | 0.237 | 37 | 17.712 | 10.359 | 5.503 | 3.73E-08 |
| Additive model | rs7161410 | 14 | 61325380 | G | A | PRKCH : Intron Variant | 0.261 | 139 | 27.367 | 48.993 | 3.91 | 9.20E-05 |
|  | rs56131196 | 19 | 44919589 | G | A | APOC1 : 500B Downstream Variant | 0.336 | 149 | 57.596 | 54.634 | 7.792 | 6.66E-15 |
|  | rs4420638 | 19 | 44919689 | A | G | APOC1 : 500B Downstream Variant | 0.337 | 150 | 58.096 | 54.884 | 7.842 | 4.44E-15 |
|  | rs12721051 | 19 | 44918903 | C | G | APOC1 : Intron Variant | 0.324 | 148 | 57.858 | 54.284 | 7.853 | 4.22E-15 |
|  | rs429358 | 19 | 44908684 | T | C | APOE : Missense Variant | 0.33 | 144 | 58.810 | 53.947 | 8.007 | 1.11E-15 |
|  | rs10414043 | 19 | 44912456 | G | A | APOC1 : 2KB Upstream Variant | 0.262 | 132 | 46.348 | 42.684 | 7.094 | 1.32E-12 |
|  | rs769449 | 19 | 44906745 | G | A | APOE : Intron Variant | 0.237 | 129 | 45.810 | 41.991 | 7.069 | 1.58E-12 |

**Supplementary Table 2.** FBAT results for rs7161410 under recessive model with and without families with K65R mutation carriers.

| **Subset** | **Marker** | **afreq** | **fam#** | **S-E(S)** | **Var(S)** | **Z** | **P** |
| --- | --- | --- | --- | --- | --- | --- | --- |
| all families | 14:61325380:G:A | 0.261 | 36 | 18.26 | 12.034 | 5.264 | 1.41E-07 |
| all families, K65R families excluded | 14:61325380:G:A | 0.259 | 35 | 17.647 | 11.284 | 5.253 | 1.49E-07 |
| NHW families | 14:61325380:G:A | 0.234 | 24 | 7.952 | 7.513 | 2.901 | 3.72E-03 |
| NHW families, K65R families excluded | 14:61325380:G:A | 0.232 | 23 | 7.339 | 6.762 | 2.822 | 4.77E-03 |
| AA families | 14:61325380:G:A | 0.459 | 2 | 0.333 | 0.444 | 0.5 | 6.17E-01 |
| Dominican families | 14:61325380:G:A | 0.333 | 10 | 9.975 | 4.077 | 4.94 | 7.81E-07 |

**Supplementary Table 3.** Number of carriers of functional variants in *PRKCH* for each genotype of rs7161410.

| rs7161410 genotype | Number of carriers of rs55645551 | | Number of carriers of rs55737090 | | Number of carriers of rs55848048 | | Number of carriers of rs2230500 | | Number of carriers of rs1884838413 | |
| --- | --- | --- | --- | --- | --- | --- | --- | --- | --- | --- |
|  | NIMH and NIA ADSP families (Total: 2247 subjects, 605 families) | NIA ADSP unrelated subjects (Total: 25660 subjects) | NIMH and NIA ADSP families (Total: 2247 subjects, 605 families) | NIA ADSP unrelated subjects (Total: 25660 subjects) | NIMH and NIA ADSP families (Total: 2247 subjects, 605 families) | NIA ADSP unrelated subjects (Total: 25660 subjects) | NIMH and NIA ADSP families (Total: 2247 subjects, 605 families) | NIA ADSP unrelated subjects (Total: 25660 subjects) | NIMH and NIA ADSP families (Total: 2247 subjects, 605 families) | NIA ADSP unrelated subjects (Total: 25660 subjects) |
| G/G (0 minor alleles) | 6 | 82 | 0 | 1 | 2 | NA | 36 | 203 | 4 | NA |
| A/G (1 minor allele) | 0 | 31 | 4 | 18 | 0 | NA | 17 | 177 | 0 | NA |
| A/A (2 minor alleles) | 0 | 0 | 4 | 9 | 0 | NA | 0 | 39 | 0 | NA |
| Missing genotype | 0 | 0 | 0 | 0 | 0 | NA | 0 | 0 | 0 | NA |

**Supplementary Figures**

**Supplementary Figure 1: Pedigree of K65R carriers in the discovery family-based dataset.** Square codes for male sibling, circle – for female sibling.

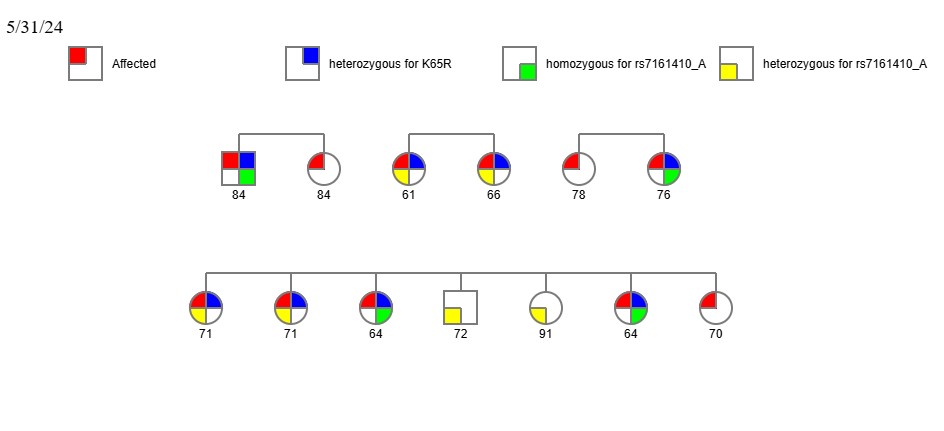

**Supplementary Figure 2: *PRKCH* expression increases in the brains of AD patients.** Violin plots of allele-specific cis-expression quantitative trait loci (eQTL) according to rs7161410 genotypes in human brain tissues based on the Genotype-Tissue Expression (GTEx, release v8) database. G and A alleles indicate the reference (major) and alternative (minor) allele types, respectively. The white line in the box plot (black) shows the median value of the expression of each genotype.

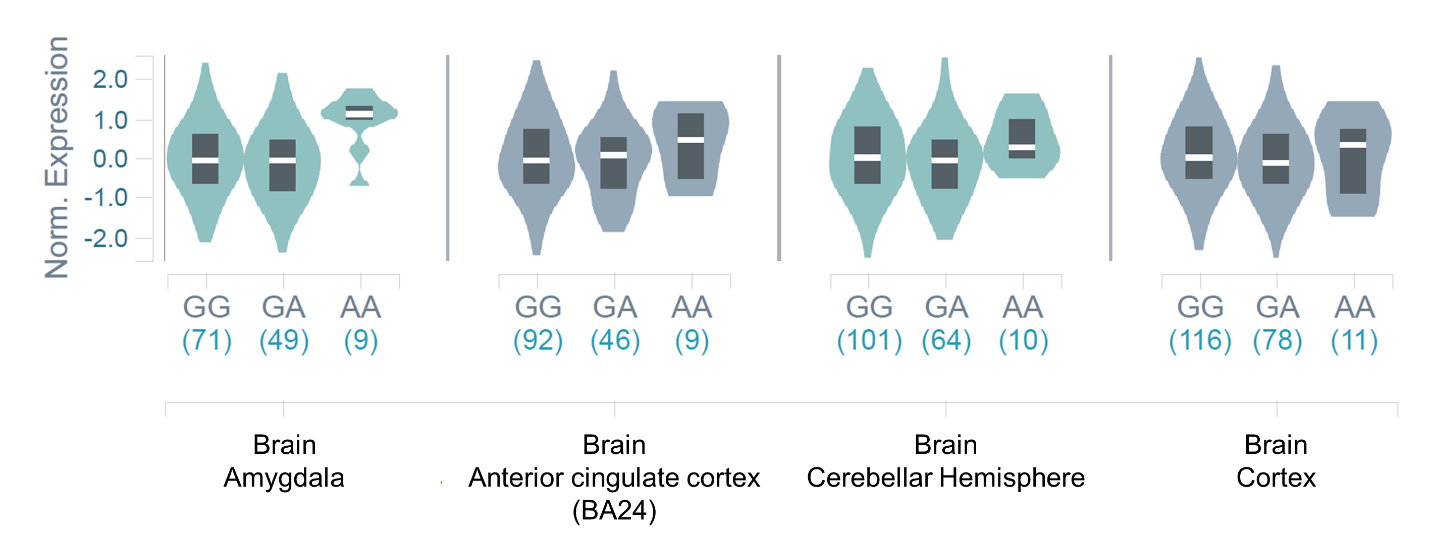

**Supplementary acknowledgements**

The All of Us Research Program is supported by the National Institutes of Health, Office of the Director: Regional Medical Centers: 1 OT2 OD026549; 1 OT2 OD026554; 1 OT2 OD026557; 1 OT2 OD026556; 1 OT2 OD026550; 1 OT2 OD 026552; 1 OT2 OD026553; 1 OT2 OD026548; 1 OT2 OD026551; 1 OT2 OD026555; IAA #: AOD 16037; Federally Qualified Health Centers: HHSN 263201600085U; Data and Research Center: 5 U2C OD023196; Biobank: 1 U24 OD023121; The Participant Center: U24 OD023176; Participant Technology Systems Center: 1 U24 OD023163; Communications and Engagement: 3 OT2 OD023205; 3 OT2 OD023206; and Community Partners: 1 OT2 OD025277; 3 OT2 OD025315; 1 OT2 OD025337; 1 OT2 OD025276. In addition, the All of Us Research Program would not be possible without the partnership of its participants.

The authors would like to thank the staff from the National Institute of Mental Health (NIMH) Divisions of Clinical and Treatment Research (DCTR) and Epidemiology and Services Research (DESR), including David Shore, MD, Mary Farmer, MD, MPH, Debra Wynne, MSW, Steven 0. Moldin, PhD, Darrell G. Kirch, MD (1989-1994), Nancy E. Maestri, PhD (1992-1994), William Huber (1989-1995), Pamela Wexler (1995-), and Darrel A. Regier, MD, MPH. They would also like to thank the study staff at all three sites and the data management staff at SRA Technologies, Inc., particularly Cheryl McDonnell, PhD, for the care and attention that they paid to all aspects of the study. The authors are also extremely grateful to the families whose participation made this work possible.

Data collection and sharing for this project was funded by the Alzheimer's Disease Neuroimaging Initiative (ADNI) (National Institutes of Health Grant U01 AG024904) and DOD ADNI (Department of Defense award number W81XWH-12-2-0012). ADNI is funded by the National Institute on Aging, the National Institute of Biomedical Imaging and Bioengineering, and through generous contributions from the following: AbbVie, Alzheimer’s Association; Alzheimer’s Drug Discovery Foundation; Araclon Biotech; BioClinica, Inc.; Biogen; Bristol-Myers Squibb Company; CereSpir, Inc.;Cogstate;Eisai Inc.; Elan Pharmaceuticals, Inc.; Eli Lilly and Company; EuroImmun; F. Hoffmann-La Roche Ltd and its affiliated company Genentech, Inc.; Fujirebio; GE Healthcare; IXICO Ltd.; Janssen Alzheimer Immunotherapy Research & Development, LLC.; Johnson & Johnson Pharmaceutical Research & Development LLC.; Lumosity; Lundbeck; Merck & Co., Inc.; Meso Scale Diagnostics, LLC.;NeuroRx Research; Neurotrack Technologies;Novartis Pharmaceuticals Corporation; Pfizer Inc.; Piramal Imaging; Servier; Takeda Pharmaceutical Company; and Transition Therapeutics.The Canadian Institutes of Health Research is providing funds to support ADNI clinical sites in Canada. Private sector contributions are facilitated by the Foundation for the National Institutes of Health (www.fnih.org). The grantee organization is the Northern California Institute for Research and Education, and the study is coordinated by the Alzheimer’s Therapeutic Research Institute at the University of Southern California. ADNI data are disseminated by the Laboratory for Neuro Imaging at the University of Southern California. A complete list of acknowledgements for the use of ADNI dataset can be found here: http://adni.loni.usc.edu/wp-content/uploads/how_to_apply/ADNI_Acknowledgement_List.pdf.

Data for this study were prepared, archived, and distributed by the National Institute on Aging Alzheimer’s Disease Data Storage Site (NIAGADS) at the University of Pennsylvania (U24-AG041689), funded by the National Institute on Aging.

The Alzheimer’s Disease Sequencing Project (ADSP) is comprised of two Alzheimer’s Disease (AD) genetics consortia and three National Human Genome Research Institute (NHGRI) funded Large Scale Sequencing and Analysis Centers (LSAC). The two AD genetics consortia are the Alzheimer’s Disease Genetics Consortium (ADGC) funded by NIA (U01 AG032984), and the Cohorts for Heart and Aging Research in Genomic Epidemiology (CHARGE) funded by NIA (R01 AG033193), the National Heart, Lung, and Blood Institute (NHLBI), other National Institute of Health (NIH) institutes and other foreign governmental and non-governmental organizations. The Discovery Phase analysis of sequence data is supported through UF1AG047133 (to Drs. Schellenberg, Farrer, Pericak-Vance, Mayeux, and Haines); U01AG049505 to Dr. Seshadri; U01AG049506 to Dr. Boerwinkle; U01AG049507 to Dr. Wijsman; and U01AG049508 to Dr. Goate and the Discovery Extension Phase analysis is supported through U01AG052411 to Dr. Goate, U01AG052410 to Dr. Pericak-Vance and U01 AG052409 to Drs. Seshadri and Fornage. Data generation and harmonization in the Follow-up Phases is supported by U54AG052427 (to Drs. Schellenberg and Wang).

The ADGC cohorts include: Adult Changes in Thought (ACT), the Alzheimer’s Disease Centers (ADC), the Chicago Health and Aging Project (CHAP), the Memory and Aging Project (MAP), Mayo Clinic (MAYO), Mayo Parkinson’s Disease controls, University of Miami, the Multi-Institutional Research in Alzheimer’s Genetic Epidemiology Study (MIRAGE), the National Cell Repository for Alzheimer’s Disease (NCRAD), the National Institute on Aging Late Onset Alzheimer's Disease Family Study (NIA-LOAD), the Religious Orders Study (ROS), the Texas Alzheimer’s Research and Care Consortium (TARC), Vanderbilt University/Case Western Reserve University (VAN/CWRU), the Washington Heights-Inwood Columbia Aging Project (WHICAP) and the Washington University Sequencing Project (WUSP), the Columbia University Hispanic- Estudio Familiar de Influencia Genetica de Alzheimer (EFIGA), the University of Toronto (UT), and Genetic Differences (GD).

The CHARGE cohorts are supported in part by National Heart, Lung, and Blood Institute (NHLBI) infrastructure grant HL105756 (Psaty), RC2HL102419 (Boerwinkle) and the neurology working group is supported by the National Institute on Aging (NIA) R01 grant AG033193. The CHARGE cohorts participating in the ADSP include the following: Austrian Stroke Prevention Study (ASPS), ASPS-Family study, and the Prospective Dementia Registry-Austria (ASPS/PRODEM-Aus), the Atherosclerosis Risk in Communities (ARIC) Study, the Cardiovascular Health Study (CHS), the Erasmus Rucphen Family Study (ERF), the Framingham Heart Study (FHS), and the Rotterdam Study (RS). ASPS is funded by the Austrian Science Fond (FWF) grant number P20545-P05 and P13180 and the Medical University of Graz. The ASPS-Fam is funded by the Austrian Science Fund (FWF) project I904),the EU Joint Programme - Neurodegenerative Disease Research (JPND) in frame of the BRIDGET project (Austria, Ministry of Science) and the Medical University of Graz and the Steiermärkische Krankenanstalten Gesellschaft. PRODEM-Austria is supported by the Austrian Research Promotion agency (FFG) (Project No. 827462) and by the Austrian National Bank (Anniversary Fund, project 15435. ARIC research is carried out as a collaborative study supported by NHLBI contracts (HHSN268201100005C, HHSN268201100006C, HHSN268201100007C, HHSN268201100008C, HHSN268201100009C, HHSN268201100010C, HHSN268201100011C, and HHSN268201100012C). Neurocognitive data in ARIC is collected by U01 2U01HL096812, 2U01HL096814, 2U01HL096899, 2U01HL096902, 2U01HL096917 from the NIH (NHLBI, NINDS, NIA and NIDCD), and with previous brain MRI examinations funded by R01-HL70825 from the NHLBI. CHS research was supported by contracts HHSN268201200036C, HHSN268200800007C, N01HC55222, N01HC85079, N01HC85080, N01HC85081, N01HC85082, N01HC85083, N01HC85086, and grants U01HL080295 and U01HL130114 from the NHLBI with additional contribution from the National Institute of Neurological Disorders and Stroke (NINDS). Additional support was provided by R01AG023629, R01AG15928, and R01AG20098 from the NIA. FHS research is supported by NHLBI contracts N01-HC-25195 and HHSN268201500001I. This study was also supported by additional grants from the NIA (R01s AG054076, AG049607 and AG033040 and NINDS (R01 NS017950). The ERF study as a part of EUROSPAN (European Special Populations Research Network) was supported by European Commission FP6 STRP grant number 018947 (LSHG-CT-2006-01947) and also received funding from the European Community's Seventh Framework Programme (FP7/2007-2013)/grant agreement HEALTH-F4-2007-201413 by the European Commission under the programme "Quality of Life and Management of the Living Resources" of 5th Framework Programme (no. QLG2-CT-2002-01254). High-throughput analysis of the ERF data was supported by a joint grant from the Netherlands Organization for Scientific Research and the Russian Foundation for Basic Research (NWO-RFBR 047.017.043). The Rotterdam Study is funded by Erasmus Medical Center and Erasmus University, Rotterdam, the Netherlands Organization for Health Research and Development (ZonMw), the Research Institute for Diseases in the Elderly (RIDE), the Ministry of Education, Culture and Science, the Ministry for Health, Welfare and Sports, the European Commission (DG XII), and the municipality of Rotterdam. Genetic data sets are also supported by the Netherlands Organization of Scientific Research NWO Investments (175.010.2005.011, 911-03-012), the Genetic Laboratory of the Department of Internal Medicine, Erasmus MC, the Research Institute for Diseases in the Elderly (014-93-015; RIDE2), and the Netherlands Genomics Initiative (NGI)/Netherlands Organization for Scientific Research (NWO) Netherlands Consortium for Healthy Aging (NCHA), project 050-060-810. All studies are grateful to their participants, faculty and staff. The content of these manuscripts is solely the responsibility of the authors and does not necessarily represent the official views of the National Institutes of Health or the U.S. Department of Health and Human Services.

The four LSACs are: the Human Genome Sequencing Center at the Baylor College of Medicine (U54 HG003273), the Broad Institute Genome Center (U54HG003067), The American Genome Center at the Uniformed Services University of the Health Sciences (U01AG057659), and the Washington University Genome Institute (U54HG003079).

Biological samples and associated phenotypic data used in primary data analyses were stored at Study Investigators institutions, and at the National Cell Repository for Alzheimer’s Disease (NCRAD, U24AG021886) at Indiana University funded by NIA. Associated Phenotypic Data used in primary and secondary data analyses were provided by Study Investigators, the NIA funded Alzheimer’s Disease Centers (ADCs), and the National Alzheimer’s Coordinating Center (NACC, U01AG016976) and the National Institute on Aging Genetics of Alzheimer’s Disease Data Storage Site (NIAGADS, U24AG041689) at the University of Pennsylvania, funded by NIA, and at the Database for Genotypes and Phenotypes (dbGaP) funded by NIH. We thank the Cure Alzheimer’s Fund for their support of this study as part of the Alzheimer’s Genome Project (RET). This research was supported in part by the Intramural Research Program of the National Institutes of health, National Library of Medicine. Contributors to the Genetic Analysis Data included Study Investigators on projects that were individually funded by NIA, and other NIH institutes, and by private U.S. organizations, or foreign governmental or nongovernmental organizations.
